# Estimating the impact of the RTS,S/AS01 malaria vaccine in Benin: A mathematical modelling study

**DOI:** 10.1101/2024.07.20.24310712

**Authors:** Sena Alohoutade, Rachel Hounsell, Codjo Dandonougbo, Rock Aikpon, Jules Degila, Sheetal Silal

**Affiliations:** Modelling and Simulation Hub, Africa, Department of Statistical Sciences, University of Cape Town, Cape Town, South Africa; Centre for Global Health Research, Nuffield Department of Medicine, Oxford University, Oxford, United Kingdom; Programme National de Lutte contre le Paludisme, Ministere de la Sante, Benin; Institut de Mathematiques et de Science Physiques, Dangbo, Benin

**Keywords:** Mathematical models, RTS, S vaccine, LLINs, SMC, Malaria modelling, Benin

## Abstract

**Background:** Malaria constitutes a major public health burden in sub-Saharan Africa. It remains a key health concern and the leading cause of death in children under five years of age in Benin. Since October 2021, the World Health Organization has recommended the use of malaria vaccines for the prevention of *Plasmodium falciparum* malaria in children living in malaria endemic areas, prioritizing areas of moderate and high transmission in sub-Saharan Africa. However, with the exception of Ghana, there is a scarcity of studies modelling the potential impact of the RTS,S/AS01 vaccine in the context of West Africa. This study addresses a gap in research by modelling the impact of malaria childhood vaccination on *Plasmodium falciparum* malaria transmission in Benin.

**Methods:** A compartmental mathematical model has been developed to estimate clinical and severe malaria cases averted in children under the age of five with the primary series (3 doses) of the RTS,S/AS01 malaria vaccine in Benin. Over a period of 10 years, scenarios including vaccine introduction at different coverage levels to supplement the use of long-lasting insecticidal nets to assess the impact of the RTS,S/AS01 vaccine on malaria transmission in Benin are modelled.

**Results:** The model projected malaria burden alleviation by malaria vaccination in Benin. The combination of childhood malaria vaccination at a coverage equivalent to the national DTP3 coverage and the current use of long-lasting insecticidal nets is projected to result in 40% reduction in malaria clinical cases and deaths among children under five years old compared to using nets alone, from 2025 to 2034. However, if the introduction of a malaria vaccine has the unintended consequence of decreased net use, cumulative benefits may be offset. A 1.5-fold decrease in the use of long-lasting insecticidal nets is projected to result in an increase in malaria burden, surpassing baseline levels despite the introduction of the vaccine.

**Conclusion:** This modelling exercise concludes that childhood vaccination is expected to avert clinical and severe cases of malaria and is an additional tool to advance malaria control efforts in Benin but potential unintended consequences of a reduction in net usage may reduce these gains.

## Introduction

Malaria constitutes a major public health burden in sub-Saharan Africa. In 2023, the World Health Organization (WHO) reported 263 million cases and 597 thousand deaths globally due to malaria. Africa accounted for 94% of these cases and 95% of the deaths, with children accounting for approximately 80% of malaria deaths in the region [1]. Two groups are particularly vulnerable to severe malaria infection: children under the age of five and expectant mothers. Children under five accounted for approximately 76% of all malaria deaths in the region [2]. Between 2015 and 2022, malaria incidence remained relatively stable. [1].

Benin is one of the fifteen countries with the highest burden of malaria; it accounted for 2% and 1.7% of malaria cases and deaths worldwide in 2023 [1]. Malaria is the leading cause of medical consultation and hospitalization in Benin. It accounts for 48% of medical consultations for children under five years old (CU5) and for 23.1% of deaths recorded in health facilities [3]. The disease exerts an economic strain on the country’s development as Beninese households allocate approximately a quarter of their yearly income to treating and preventing malaria [4]. Despite extensive attempts to control and eliminate the disease, malaria continues to have a substantial impact on the health and well-being of millions of people across Africa.

Malaria preventive interventions include insecticide residual spraying, the use of bed nets and prophylactic treatment of pregnant women, infants, and children [5]. However, these interventions are not exempt from challenges posed by insecticide-resistance vectors and drug-resistance parasites. Therefore, among the various strategies employed to tackle malaria, vaccination has emerged as a promising approach to complement existing preventive and treatment measures.

RTS,S/AS01_E_ (RTS,S), is one of two candidate vaccines recommended by the WHO in October 2021. It took over 30 years to be developed, approved, and to enter pilot implementation studies [6]. In a pivotal Phase III trial, four doses of RTS,S administered to children of five months of age or older resulted in a 39% reduction in clinical malaria and a 29% reduction in severe malaria, sustained over a four-year follow-up period [7]. Following this trial, RTS,S underwent evaluation in Ghana, Malawi, and Kenya as part of a widespread WHO pilot program [8]. From October 2021, the WHO recommended RTS,S for use in children in sub-Saharan Africa and other high burden areas where *Plasmodium falciparum* transmission is moderate to high [9].

In April 2024, Benin introduced the RTS,S malaria vaccine into its Expanded Programme on Immunization (EPI), marking a significant milestone in the country’s malaria control efforts. This followed Benin’s strong engagement with Gavi, the Vaccine Alliance, through which it received a total of 215,900 RTS,S doses to support the rollout of the vaccine [10]. In the early phases of this rollout, there remains a need to model the impact and cost effectiveness of vaccination to inform its implementation in Benin and in countries that have shown interest in its introduction.

Mathematical models of malaria transmission can provide evidence to help inform a variety of decision-making processes such as identifying potentially beneficial combinations of interventions, establishing attainable coverage targets, anticipating the impact of new interventions, and evaluating the risk of malaria resurgence [11]. Since the recommendation of RTS,S by the WHO, many modelling studies have evaluated its impact in sub-Saharan Africa [6,12,13].

A WHO working group used four mathematical models to assess the health impact and cost effectiveness of the RTS,S vaccine in an African setting [14–17]. Following this, Penny et al.[18] conducted a comparative analysis of the four malaria models and predicted the public health impact of the RTS,S malaria vaccine in a wide range of African settings in 2016. However, LMIC countries’ health-care systems, vaccine schedules, and cost assumptions differ greatly. Recognizing this, national policymakers are increasingly seeking evidence from within their own countries to inform their decisions [12]. Among the studies that have modelled malaria vaccine impact in sub-Saharan Africa, Hogan et al.[19], found that in high-endemic areas, adopting vaccine coverage similar to that of the third dose of diphtheria, tetanus, and pertussis (DTP3), could prevent 4.3 million malaria cases and 22,000 deaths in CU5 each year across sub-Saharan Africa.

There is a scarcity of studies modelling the potential impact of RTS,S in the context of West Africa and only a few of the RTS,S vaccine modelling studies evaluated the vaccine’s impact taking into account existing interventions, such as long-lasting insecticidal nets (LLINs) [20]. This may be attributed to insufficient resources to support malaria vaccination campaigns, as there is a gap between the funding required and the resources available for malaria control efforts [21,22]. This may also be attributed to the limited availability of accurate, region-specific data on malaria transmission dynamics. This limitation arises from resource constraints, gaps in surveillance systems, and delays in data collection and analysis, which collectively contribute to the relatively low number of research publications emerging from the West African region, with a lower number lead by West African researchers [23].

In this study, a compartmental model is employed to project the relative reduction in clinical and severe malaria cases achieved by introducing the RTS,S malaria vaccine in relation to the distribution and use of LLINs in Benin. This will not only provide an assessment of the additive nature of the two interventions but provide insight into the impact of social behavioural change communication and vaccination campaigns.

### Study setting and data

The Republic of Benin is located in West Africa with a population estimated to be approximately 14.6 million in 2024. In Benin, malaria incidence is 150 per 1,000 population at risk and the death rate in CU5 is 96 per 1,000 [24]. Malaria transmission is variable across Benin with seasonal and geographic fluctuations closely correlated with rainfall patterns, climate, and topography. The geography and climate provide a favourable environment for malaria persistence [3].

Several interventions have been implemented to prevent malaria in Benin. These interventions include the use of LLINs, seasonal malaria chemoprevention (SMC) and intermittent preventive treatment of malaria during pregnancy (IPTp).

The primary pillar of Benin’s vector control strategy is LLINs [25]. With funding from President’s Malaria Initiative (PMI), Benin began SMC in two health districts in 2019. Following positive outcomes, such as a coverage of 97% of the target group of children and a decrease in morbidity, SMC was expanded to 15 out of the 34 eligible health zones in 2021– 2023 [26].

### Data description

Population data were sourced from the United Nations World Population Prospects, facilitating projections of population size, growth, and estimated numbers of individuals for each year of life, relying on fertility and mortality data. We used country-specific coverage for DTP3 vaccination from the WHO/UNICEF Estimates of National Immunization Coverage (WUENIC) as a proxy for RTS,S vaccine coverage [27]. Estimated annual clinical malaria cases, deaths and historical LLINs coverage estimates for Benin were obtained from the World Malaria Report (WMR) annexes [1].

## Methods

### Description of the model

The model is a deterministic age-structured dynamic compartmental model accounting for LLIN use and seasonality to estimate the impact of the RTS,S malaria vaccine on clinical malaria cases and deaths in Benin.

As the burden of malaria varies by age, the model considered the age structure of the population, determined by both the vaccination schedule and the age profile for the severity of the disease. According to a large phase III trial that included several endemic areas of Africa, the RTS,S vaccine efficacy against clinical malaria begins at 74% in children aged 5 to 17 months, a few weeks after the last immunization, and drops to 9% after 5 years [28].

In Ghana, the vaccine is administered to children in four doses, beginning at six months, followed by doses at 7, 9, and 18 months [29]. Following the 215,900 doses of the malaria vaccine acquired in January 2024, Benin has begun administering three doses to children at 6, 7, 9 months and a fourth dose as a booster for children under 24 months of age [10]. Considering potential constraints (e.g., financial constraints, loss to follow-up), here we assess the impact of only three doses. We assumed that children received the three doses at 6, 7, and 9 months.

The model categorizes the human population into three age groups: children from birth to six months, those aged six months to five years, and individuals aged five years and older (Fig 1). Susceptible individuals are exposed to infectious bites at a rate depending on mosquito density and infectivity. Following exposure (E_i_), there is a latent period after which the infected individuals may experience an asymptomatic (A_i_) episode of malaria according to their immunity level or develop clinical symptoms (C_i_). The chance of symptomatic disease decreases with continued exposure due to the development of naturally acquired immunity. Asymptomatic individuals do not display symptoms but can transmit parasites to mosquitoes. However, asymptomatic individuals recover naturally after a period of time. A clinical episode, where individuals are symptomatic, can be uncomplicated (C_i_) or progress to a severe malaria episode (F_i_), which can result in death.

**Fig 1:**
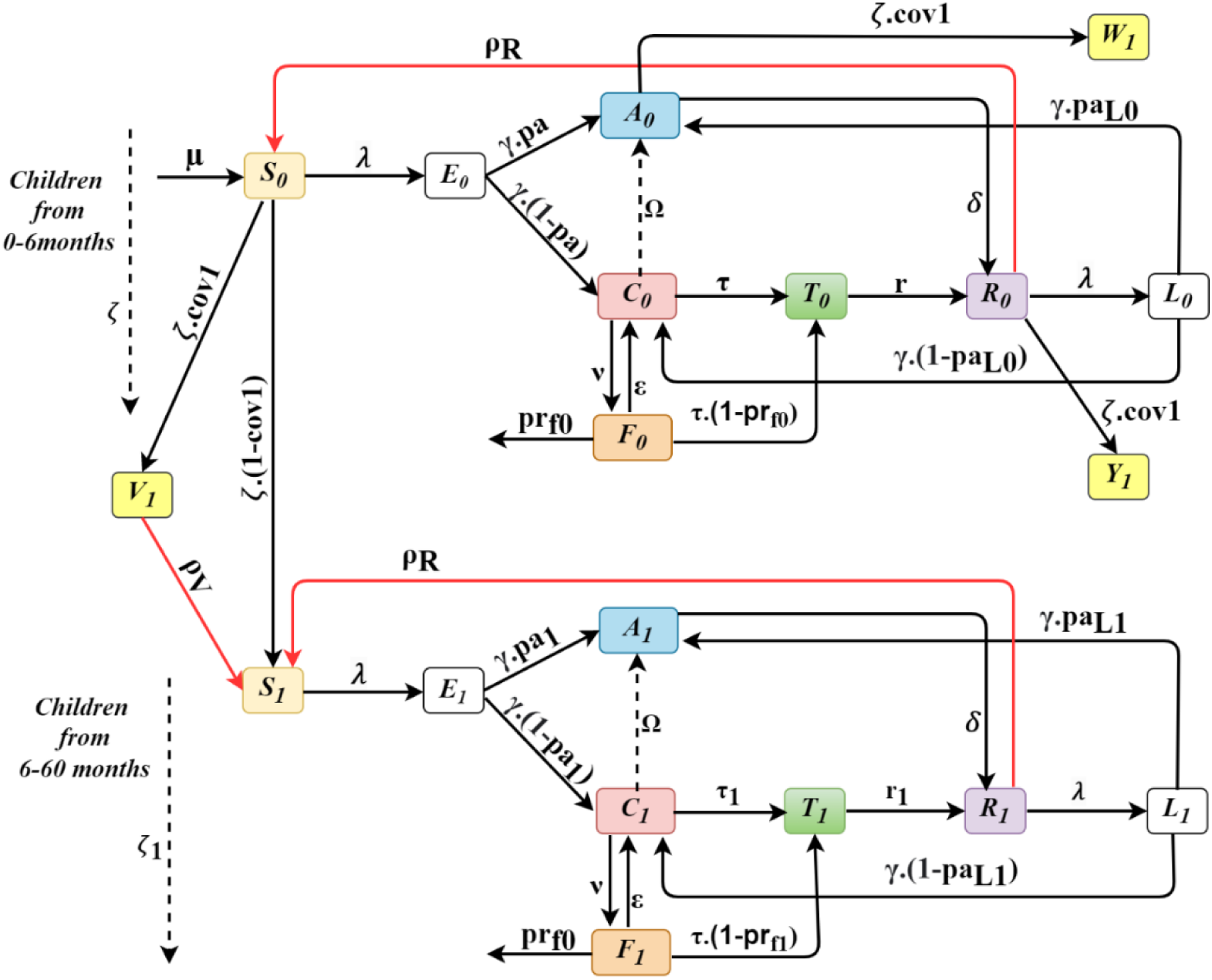
Compartmental model showing the first two age groups with compartments Si (Susceptible), E_i_ (Exposed), A_i_ (Asymptomatic), C_i_ (Symptomatic), T_i_ (Treated), R_i_ (Recovered), L_i_ (Second Exposure) and V_i_, W_i_,Y_i_ (Vaccination compartments).

We consider a natural mortality rate from each compartment. Additionally, malaria-related directly-attributable deaths are explicitly considered in the severe stage of modelled infection. We assume that children age between groups at a rate 𝜁.

We also assume that susceptible, asymptomatic or recovered individuals at the age of six months may be vaccinated. Vaccinated individuals in the model may be infected and develop an asymptomatic or symptomatic episode of malaria. Vaccinated individuals lose immunity due to waning after four years. LLINs coverage and use and routine treatment for malaria are also included in the model.

Figure 1 shows a diagram of the compartmental model showing the first two age groups. Seasonality is modelled by multiplying the force of infection of the model by a seasonality factor. The average monthly rainfall at a country level was standardized and fitted to a trigonometric function, which was then used as a forcing function to account for seasonality. The full description of the model, parameters table and differential equations are available in the supplementary file.

### Scenarios

Six scenarios of vaccine coverage and effective LLIN coverage, classified under two categories, are simulated over a period of 10 years to provide estimates of the malaria burden alleviated by these interventions in CU5.

The baseline represents the existing effective LLIN coverage, assuming that LLIN coverage and use will remain at the average levels for 2020–22 [30]. It serves as a comparison for the other scenarios.

The first three scenarios model vaccine introduction with no impact on other interventions. The first scenario (76% vaccination + 43.2% effective LLIN coverage) includes the introduction of vaccination at DTP3 coverage level (76%) with no impact on net use and distribution. In this scenario, all eligible children are included in the target population; however, the uptake of the vaccine is assumed based on the administration of the third dose of DTP3, using country-level 2022 DTP3 coverage data from the WUENIC [31].

The second scenario (50% vaccination + 43.2% effective LLIN coverage) includes vaccine introduction assuming that a lower level than DTP3 coverage is achieved (50%) with no impact on net use and distribution.

The third scenario (85% vaccination + 43.2% effective LLIN coverage) includes vaccine introduction, assuming a coverage level higher than DTP3 is achieved (85%) with no impact on net use and distribution.

The second three scenarios models vaccine introduction with impact on effective LLIN coverage. The fourth scenario (76% vaccination + 28.8% effective LLIN coverage), includes vaccine introduction at DTP3 coverage with a negative impact on effective LLIN coverage. It assumes a vaccine coverage of 76% in children (as in Scenario 1) and a decrease to 28.8% in effective LLIN coverage (from the baseline of 43.2%) to simulate the potential negative impact of vaccine introduction on effective LLIN coverage. This scenario considers the possibility that the perceived protection from the vaccine may lead to a reduction in the use of LLINs among the population.

In the fifth scenario (85% vaccination + 50.4% effective LLIN coverage), we considered a social behavioural change communication campaign (SBCC) to improve vaccine coverage and net use. The campaign is modelled to result in vaccine coverage of 85% in the targeted population, and an increase in effective LLIN coverage from baseline (43.2%) to 50.4%.

The sixth scenario (0% vaccination + 28.8% effective LLIN coverage) is considered a reverse scenario, designed to assess the relative contribution of vaccination by comparing outcomes with and without its addition under reduced effective LLIN coverage. It assumes no vaccine introduction (0% coverage in children) and a decline in effective LLIN coverage from 43.2% (baseline) to 28.8%. The reduction in effective LLIN coverage may reflect either a decrease in net availability due to funding constraints or reduced use resulting from behavioural changes within the population. This scenario highlights the relative contribution of vaccination by comparing outcomes with and without its introduction under reduced effective LLIN coverage.

Table 1 presents an overview of the scenarios.

**Table 1.** Scenarios and coverage levels for modelling.

|  |  | Scenarios |  |  |  |
| --- | --- | --- | --- | --- | --- |
| Categories | Variables | Baseline | Scenario 1<br>(DTP3_cov) | Scenario 2<br>(Low_cov) | Scenario 3<br>(High_cov) |
| Vaccine Introduction<br>With no impact on other interventions | Vaccine coverage | - | 76% | 50% | 85% |
|  | Effective LLIN coverage | 2020–22 effective coverage<br>43.2% (72% coverage *60% use) |  |  |  |
|  | Variables |  | Scenario 4<br>(DTP3_cov+low LLIN) | Scenario 5<br>Optimistic<br>(High_cov+high LLIN) | Scenario 6<br>Reduced LLIN |
| Vaccine introduction with impact on other interventions | Vaccine coverage |  | 76% | 85% | - |
|  | Effective LLIN coverage |  | 28.8%<br>(72% coverage *40% use) | 50.4%<br>(72% coverage *70% use) | 28.8%<br>(72% coverage *40% use) |

### Sensitivity analysis

A sensitivity analysis was conducted to evaluate the validity of our results and determine how changes in key parameters influence the projected impact on clinical malaria cases over the 10-year period. The parameters with the largest impact on clinical malaria cases were LLIN use, treatment seeking period, vaccine coverage and vaccine waning. The full detail on the sensitivity analysis conducted is available in the supplementary file.

### Estimation of uncertainty intervals for model outputs

Uncertainty intervals were estimated by running 200 simulations per model scenario. In each simulation, parameters were randomly sampled from their respective distributions within defined lower and upper bounds (Table S2 in the supplementary file). Daily malaria case estimates were then aggregated across all simulations, and the median, along with the 5th and 95th quantiles, was computed for each day.

## Results

Six scenarios, each simulated over a ten-year period (2025-2034), were used to evaluate the impact of varying vaccine coverage levels and effective LLIN coverage on malaria cases and deaths in CU5 and the total population in Benin. The reduction in malaria burden associated with each scenario over the cumulative period from 2025 to 2034 was assessed. This approach highlights the broader impact of vaccination by comparing the projected cases, severe cases, and deaths averted under each scenario to the baseline estimates.

Figures 2 and 3 illustrate the projected trajectories of daily clinical cases in CU5 for Scenarios 1-3 and 4-6 respectively. Scenarios 1-3 simulate vaccine introduction at different coverages with no impact on effective LLIN coverage while, scenarios 4-6 simulate vaccine introduction at different coverages with impact on effective LLIN coverage.

**Fig 2:**
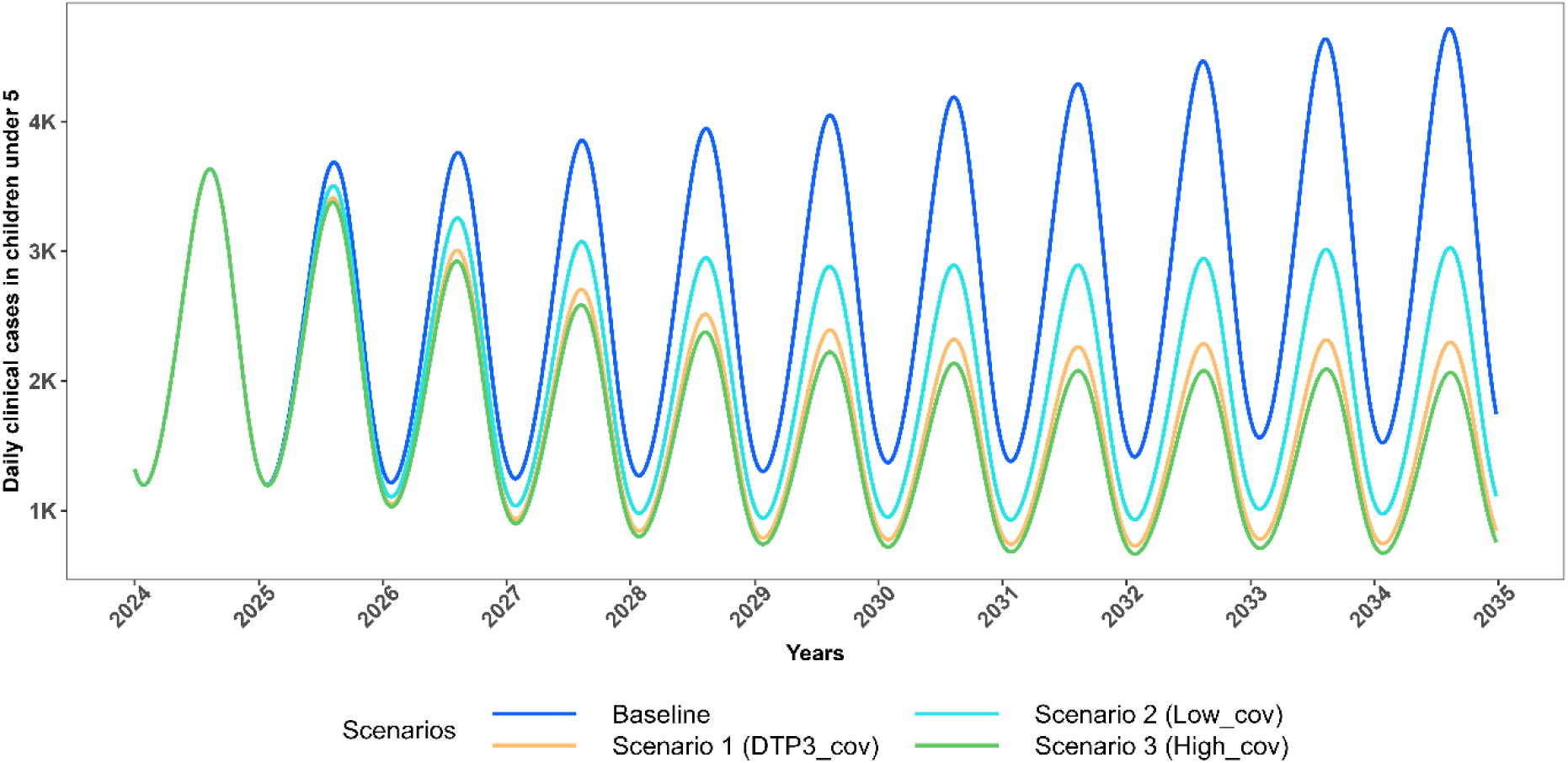
Projected trajectories of *P.falciparum* malaria daily clinical cases in CU5 for scenarios with no change in effective LLIN coverage (2025-2034) Trends in projected daily clinical malaria cases among children under five (CU5) from 2024 to 2035, comparing intervention scenarios assuming no change in effective LLIN coverage (maintained at 43.2%). The Baseline scenario models 0% vaccination coverage (blue). Scenario 1 assumes 76% vaccination coverage with 43.2% effective LLIN coverage (orange); Scenario 2 assumes 50% vaccination coverage with 43.2% effective LLIN coverage (cyan); and Scenario 3 assumes 85% vaccination coverage with 43.2% effective LLIN coverage (green).

**Fig 3:**
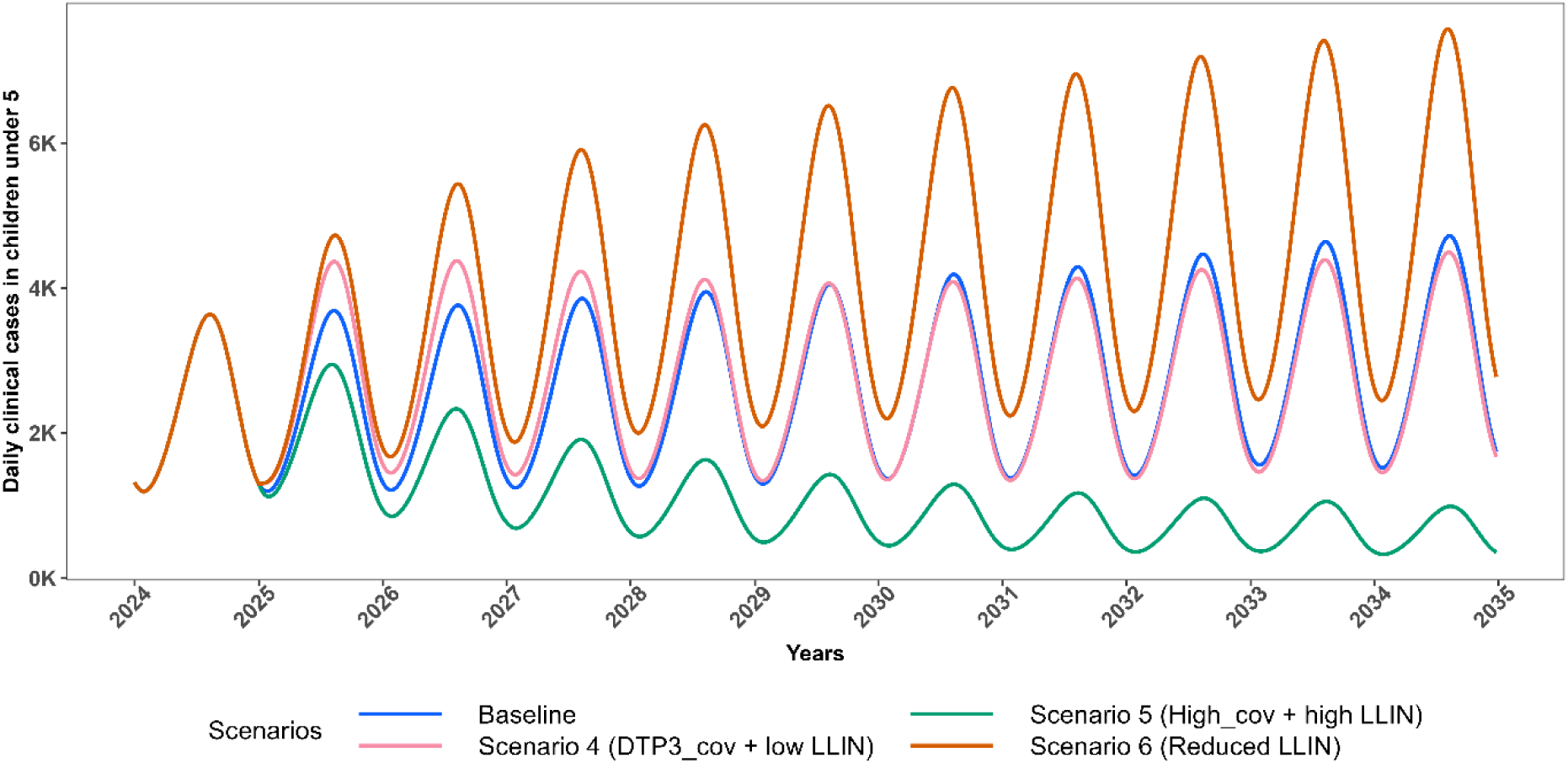
Projected trajectories of *P.falciparum* malaria daily clinical cases in CU5 for scenarios with change in effective LLIN coverage (2025-2034) Trends in projected daily clinical malaria cases among children under five (CU5) from 2024 to 2035, comparing intervention scenarios involving changes in effective LLIN coverage to the baseline. The Baseline scenario assumes 0% vaccination coverage with 43.2% effective LLIN coverage (blue). Scenario 4 assumes 76% vaccination coverage with 28.8% effective LLIN coverage (pink); Scenario 5 assumes 85% vaccination coverage with 50.4% effective LLIN coverage (dark green); and Scenario 6 assumes 0% vaccination coverage with 28.8% effective LLIN coverage (dark orange).

In the baseline scenario, the daily incidence of clinical malaria increases with population growth over time though incidence rate remains relatively stable throughout the simulation period (2025-2034). In Scenario 1, which simulates the introduction of RTS,S vaccine at DTP3 coverage levels, there is a substantial reduction of 40% in clinical malaria cases in CU5 over ten years, equivalent a 1.67-fold reduction in all outcomes compared to the baseline (Table 2-3, Figures 6-7, S6).

**Table 2:** Projected number of cases and deaths in CU5 across the six scenarios compared to the baseline.

| **In children**<br>**under 5 years** | **Number of cases** |  | **Number of deaths** |  |
| --- | --- | --- | --- | --- |
|  | **By 2029** | **By 2034** | **By 2029** | **By 2034** |
| **Baseline** | 4,330,100 | 9,505,730 | 30,600 | 67,500 |
| **Scenario 1** | 3,124,700 | 5,767,500 | 22,000 | 40,700 |
| **Scenario 2** | 3,504,500 | 6,905,100 | 24,700 | 48,800 |
| **Scenario 3** | 3,000,900 | 5,408,500 | 21,000 | 38,000 |
| **Scenario 4** | 4,863,900 | 9,889,940 | 34,500 | 69,900 |
| **Scenario 5** | 2,272,400 | 3,647,300 | 15,900 | 25,600 |
| **Scenario 6** | 6,676,000 | 15,171,400 | 47,500 | 108,100 |

**Table 3:** Projected Changes in Percentage of Cases and Deaths in CU5 across the six scenarios compared to the baseline.

| **In children**<br>**under 5 years** | **Percentage of cases** |  | **Percentage of deaths** |  |
| --- | --- | --- | --- | --- |
|  | **By 2029** | **By 2034** | **By 2029** | **By 2034** |
| **Scenario 1** | -30% | -40% | -28% | -41% |
| **Scenario 2** | -19% | -27% | -19% | -28% |
| **Scenario 3** | -30% | -45% | -30% | -46% |
| **Scenario 4** | +11% | +4% | +10% | +4% |
| **Scenario 5** | -47% | -62% | -46% | -62% |
| **Scenario 6** | +33% | +38% | +32% | +39% |

*Positive values show an increase compared to the baseline and negative values show a decrease compared to the baseline.*

Scenario 2, which simulates the introduction of the vaccine at coverage levels lower than DTP3 (50% vs 76% coverage), projected a 27% reduction in clinical cases, severe cases, and deaths in CU5 —less than the impact observed in Scenario 1 (Table 2-3, Figures 6-7, S6). This corresponds to a 1.37-fold reduction in all outcomes over ten years. This comparison highlights that a decrease in vaccine coverage, as seen in Scenario 2, undermines the potential health benefits, reinforcing the need to maintain or even enhance vaccine coverage to maximize public health gains.

Scenario 3 simulates the introduction of the vaccine, assuming a coverage level higher than DTP3 is achieved (85% vs 76% coverage) with no impact on effective LLIN coverage. Under this scenario, there is a substantial reduction projected of 45% in clinical malaria cases in CU5 (Table 2-3, Figures 6-7, S6). This highlights the additional health benefits of increasing vaccine coverage, reinforcing the importance of achieving high uptake to maximize malaria burden reduction. The trajectory of *P. falciparum* malaria clinical cases in CU5 for each scenario influences the corresponding trends in severe cases and malaria-related deaths in CU5 (See Supplementary file S3-S4).

Scenarios 4 to 6 were constructed to explore the possible unintended impact of vaccine introduction on other interventions. Scenario 4 simulates vaccine introduction at DTP3 coverage levels combined with a reduction in effective LLIN coverage (from 43.2% to 28.8%). Initially (2025–2029), daily clinical malaria cases rise compared to the baseline in CU5, but from 2030 onward, cases decline (Fig 3). This pattern reflects the vaccine’s age-specific administration (6 months to 2 years). Initially, the impact is offset by increased malaria cases in older unvaccinated children (aged 2 to 5 years), resulting from a reduction in effective LLIN coverage. However, by 2030, most under-fives are vaccinated, leading to a significant reduction in clinical cases. This scenario projected approximately 4% increase in all outcomes compared to the baseline in CU5 over the 10 year period (Table 2-3, Figures 6-7, S6). Notably, this scenario substantially impacts the total population, resulting in an approximate 20% increase in malaria cases and a 12% rise in malaria-related deaths compared to baseline levels (Figure 8, S7).

This finding indicates that the introduction of vaccination at DTP3 coverage levels, alongside reduced effective LLIN coverage, may mask the vaccine’s impact in CU5, as this scenario closely resembles the baseline over the ten-year period. Overall, the increase in malaria cases is more pronounced in the total population than in CU5, likely because LLINs serve as the main intervention for protection across the entire population, and Scenario 4 involved a reduction in their effective coverage. This highlights the critical role of sustained LLIN distribution and use in maximizing vaccination benefits.

Figures 4 and 5 respectively illustrate the projected trajectories of daily clinical cases in the total population for each set of the scenarios.

**Fig 4:**
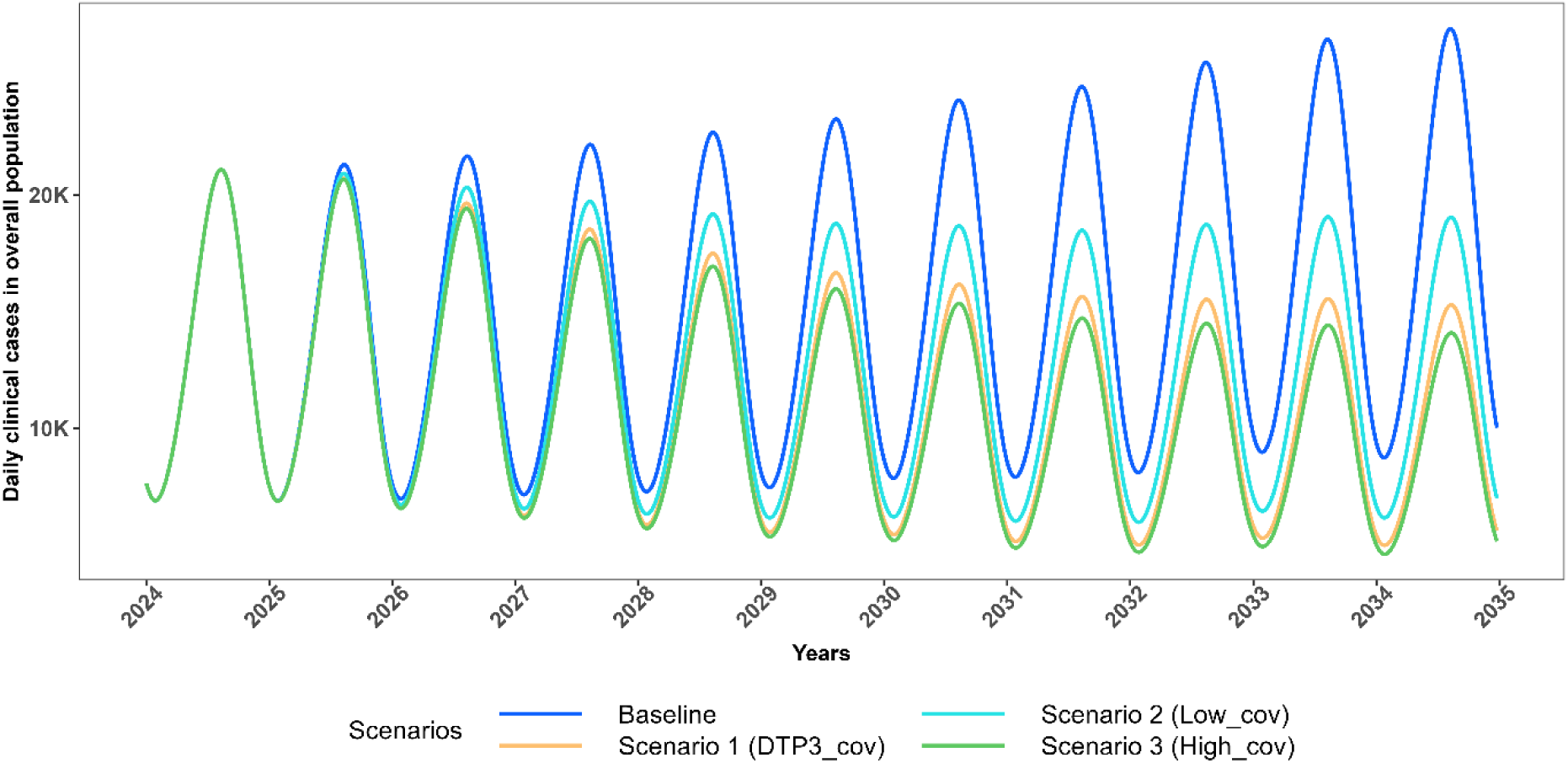
Projected trajectories of *P.falciparum* malaria daily clinical cases in the total population for scenarios with no change in effective LLIN coverage (2025-2034) Trends in daily clinical malaria cases in total population from 2024 to 2035, comparing intervention scenarios assuming no change in effective LLIN coverage (maintained at 43.2%). The Baseline scenario models 0% vaccination coverage (blue). Scenario 1 assumes 76% vaccination coverage with 43.2% effective LLIN coverage (orange); Scenario 2 assumes 50% vaccination coverage with 43.2% effective LLIN coverage (cyan); and Scenario 3 assumes 85% vaccination coverage with 43.2% effective LLIN coverage (green).

**Fig 5:**
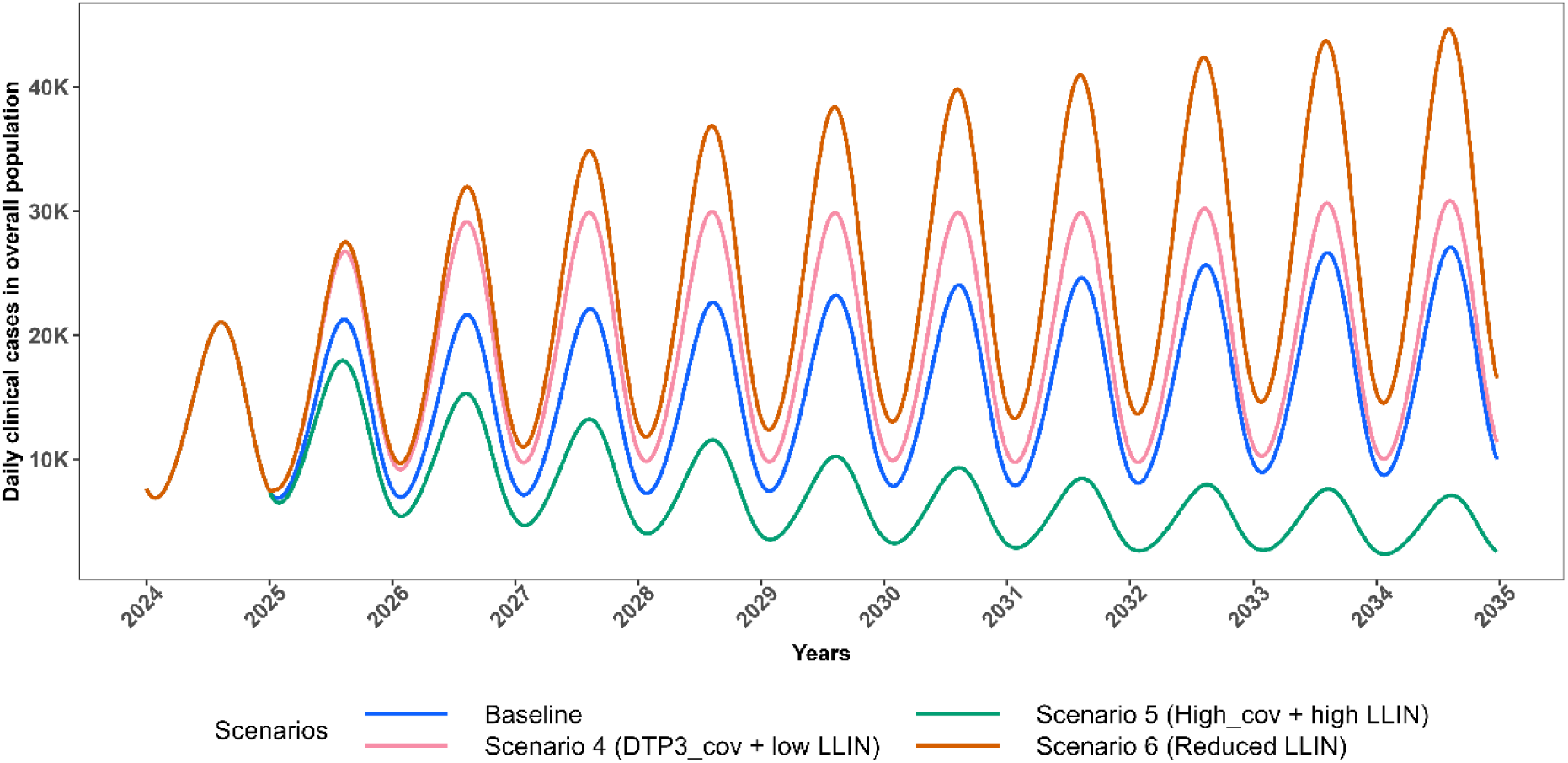
Projected trajectories of *P.falciparum* malaria daily clinical cases in the total population for scenarios with change in effective LLIN coverage (2025-2034) Trends in projected daily clinical malaria cases in total population from 2024 to 2035, comparing intervention scenarios involving changes in effective LLIN coverage to the baseline. The Baseline scenario assumes 0% vaccination coverage with 43.2% effective LLIN coverage (blue). Scenario 4 assumes 76% vaccination coverage with 28.8% effective LLIN coverage (pink); Scenario 5 assumes 85% vaccination coverage with 50.4% effective LLIN coverage (dark green); and Scenario 6 assumes 0% vaccination coverage with 28.8% effective LLIN coverage (dark orange).

In Scenario 5, which simulates the improvement of vaccine coverage and increase in effective LLIN coverage through SBCC, there is a substantial reduction of approximately 62% in clinical malaria cases in CU5 compared to the baseline, corresponding to a 2.86-fold reduction in all outcomes over the 10-year period. This reduction underscores the added benefit of enhanced intervention coverage, as Scenario 5 (85% vaccination + 50.4 % effective LLIN coverage) has the potential to achieve approximately 1.7 times the reduction observed with Scenario 1 (76% vaccination + 43.2 % effective LLIN coverage) in CU5. Moreover, Scenario 5 projected more cases and deaths averted across the entire population, attributable to an increase in effective LLIN coverage (Table 2-3, Figures 6-7, S6).

**Fig 6:**
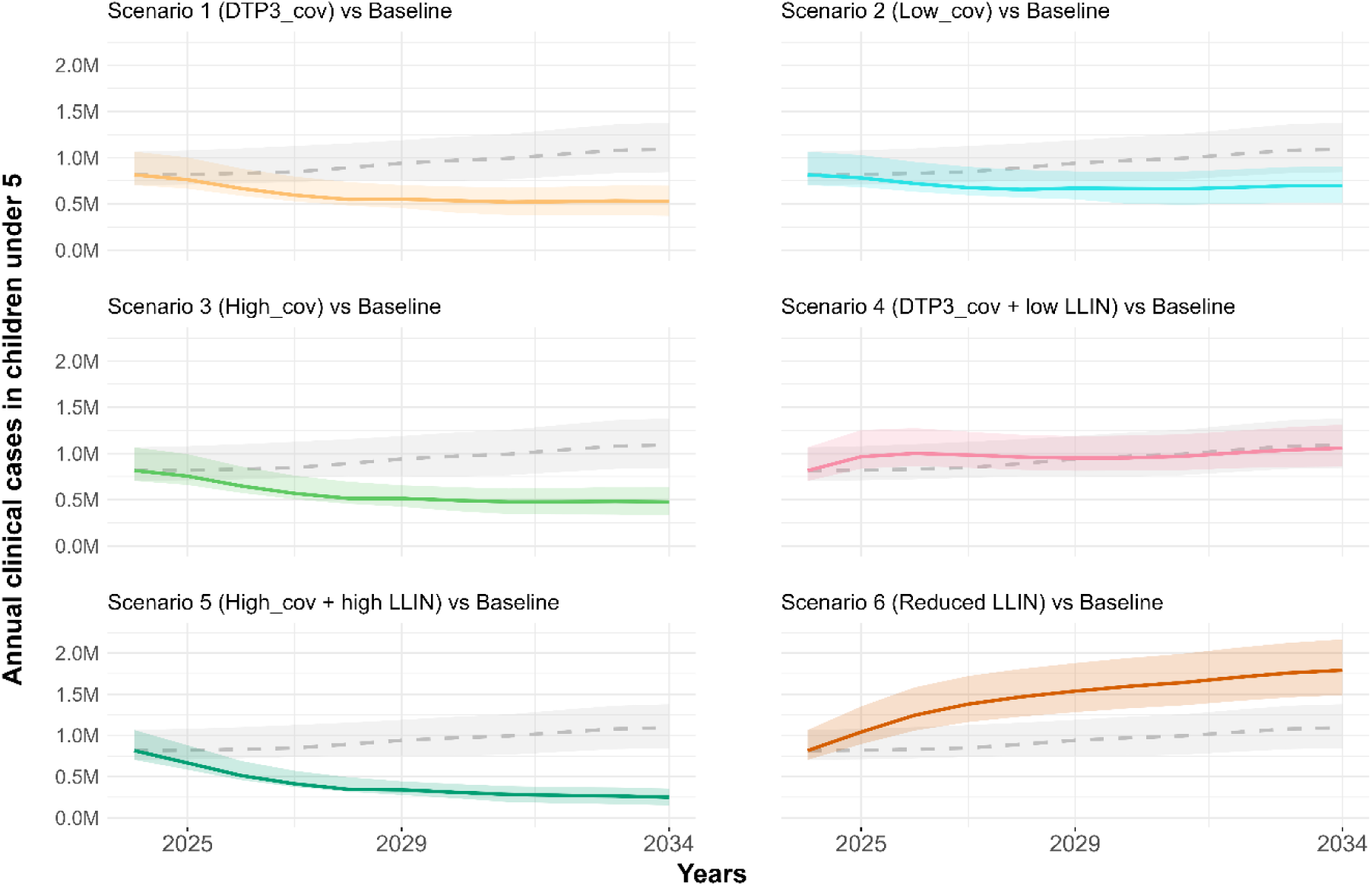
Comparison of annual clinical malaria cases projections under six scenarios with the baseline (2024–2034). The baseline is represented by a dashed grey line with a shaded grey confidence interval, indicating the range of uncertainty (90% uncertainty interval). Scenario 1 (orange), Scenario 2 (cyan), Scenario 3 (green), Scenario 4 (pink), Scenario 5 (dark green), and Scenario 6 (dark orange) show projections under different intervention assumptions. The Baseline scenario assumes 0% vaccination coverage with 43.2% effective LLIN coverage. Scenario 1 assumes 76% vaccination coverage with 43.2% effective LLIN coverage; Scenario 2 assumes 50% vaccination coverage with 43.2% LLIN coverage; and Scenario 3 assumes 85% vaccination coverage with 43.2% effective LLIN coverage. Scenario 4 assumes 76% vaccination coverage with 28.8% effective LLIN coverage; Scenario 5 assumes 85% vaccination coverage with 50.4% LLIN coverage; and Scenario 6 assumes 0% vaccination coverage with 28.8% effective LLIN coverage. The solid lines represent the median estimates, while the shaded areas indicate the 90% uncertainty intervals (5th and 95th percentiles).

**Fig 7:**
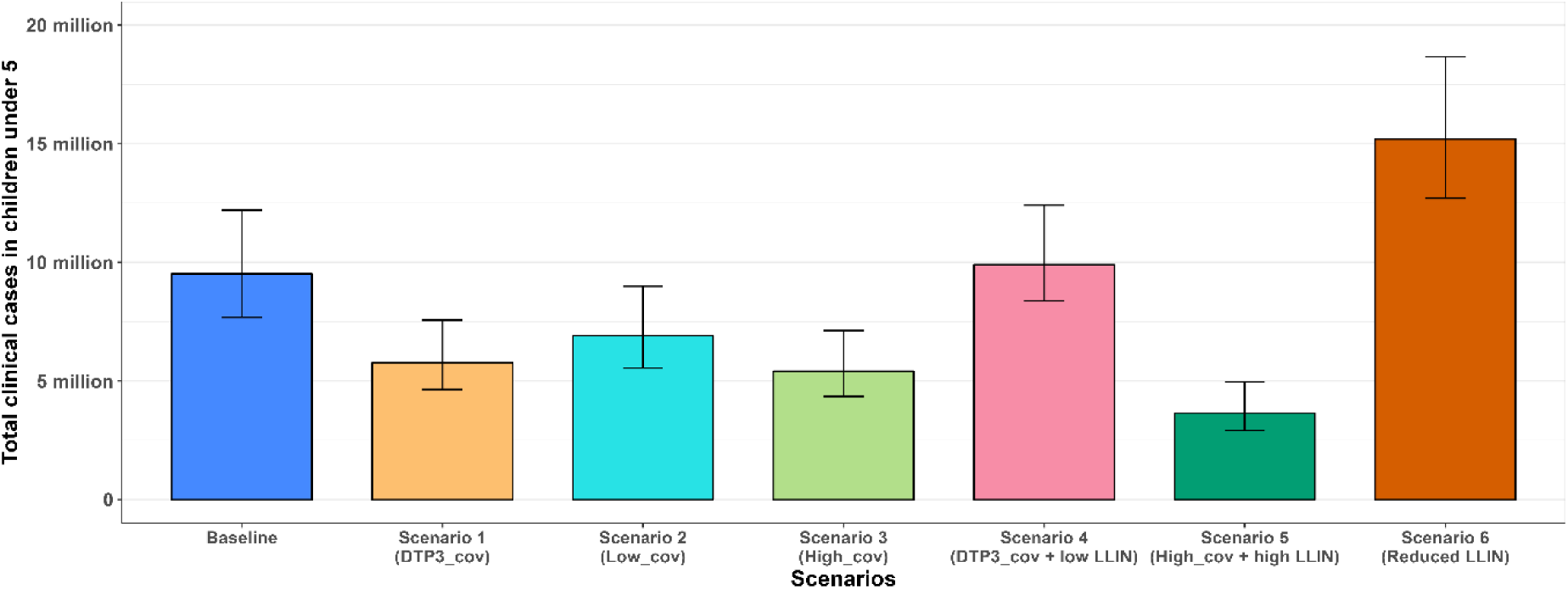
Total projected clinical cases in CU5 over 10 years (2025-2034) Bar graph showing the total malaria clinical cases in CU5 for the baseline (blue) and six other scenarios: Scenario 1 (orange), Scenario 2 (cyan), Scenario 3 (green), and Scenario 4 (pink), Scenario 5 (dark green), and Scenario 6 (dark orange). The bars represent the 90% uncertainty intervals (5th and 95th percentiles) for each scenario.

Scenario 6 detailing no vaccine introduction with a reduction in effective LLIN coverage (from 43.2% to 28.8%), projects an increase in clinical malaria cases, surpassing baseline levels in both CU5 and the total population. This scenario is designed to assess the relative contribution of vaccination by comparing outcomes with and without its addition under reduced effective LLIN coverage. Under this scenario, there is a substantial 38% increase in clinical malaria cases in CU5 (Table 2-3, Figures 6-7, S6). This emphasizes that reducing effective LLIN coverage without introducing vaccination can lead to a worsening malaria burden as Scenario 6 has the potential to result in a large increase in malaria cases (38%), particularly in CU5 compared to Scenario 4 which results in only 4% increase due to the result that the vaccine benefits are partially masked by the reduced effective LLIN coverage.

This underlines the critical importance of both vaccination and LLINs in malaria control.

Figures 6 and 7 respectively illustrate the projected trajectories of annual clinical cases over time for all scenarios and the total projected clinical cases in CU5. Additionally, Figure 8 illustrates the total projected clinical cases in the total population.

**Fig 8:**
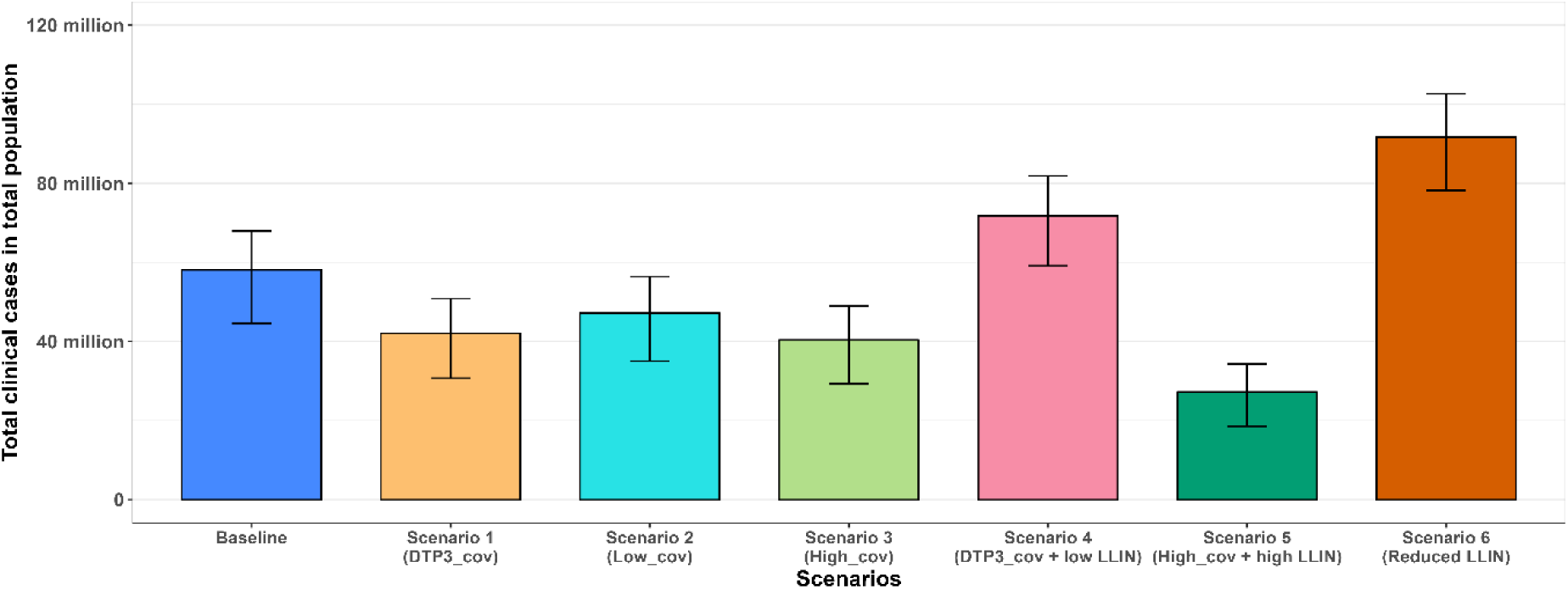
Total projected cases in the total population over 10 years (2025-2034) Bar graph showing the total clinical cases in the total for the baseline (blue) and six other scenarios: Scenario 1 (orange), Scenario 2 (cyan), Scenario 3 (green), and Scenario 4 (pink), Scenario 5 (dark green), and Scenario 6 (dark orange). The bars represent the 90% uncertainty intervals (5th and 95th percentiles) for each scenario.

## Discussion

Based on a mathematical compartmental model of *P. falciparum* malaria, our study estimated the clinical cases and deaths averted in Benin, West Africa, following the introduction of the RTS,S malaria vaccine over a 10-year period (2025-2034). Considering six scenarios for vaccine coverage and LLIN use, this study predicts that vaccine coverage at a national level of 85% in children, access to LLINs at 2020-2022 coverage levels and an increased use of LLINs at 70% across the country will result in a 62% reduction in clinical malaria cases and deaths in children under five years of age. However, the introduction of the RTS,S malaria vaccine at DTP3 coverage levels (76%) to supplement the current use of nets (60%) was also predicted to result in a substantial 40% reduction in clinical malaria cases and deaths in children under five. This suggests that vaccination can play a crucial role in reducing the malaria burden in children under five, even when LLIN use remains at current levels.

Our results qualitatively align with those reported in a modelling study by Hogan et al. [19], which estimated the impact of the RTS,S malaria vaccine allocation strategies in sub-Saharan Africa. The study revealed that in high-endemic areas, adopting vaccine coverage similar to that of DTP3 could prevent 4.3 million malaria cases and 22,000 deaths in children under five each year in sub-Saharan Africa. Moreover, our results qualitatively align with the study conducted by Hamilton et al. [6], which indicated that the widespread implementation of a malaria vaccine has the potential to decrease the malaria burden in Africa.

Scenario 4 was included in our simulation to project the impact of a decrease in net use on malaria transmission. When considered additively, the combination of vaccination and LLINs is predicted to be effective in reducing malaria clinical cases. However, factors affecting their implementation must also be considered. The introduction of a malaria vaccine may negatively affect net use, which could lead to a relatively smaller reduction (or even an increase) in malaria burden [6]. In Scenario 4, the model predicts that a 14.32% national decline in effective LLIN coverage compared to current levels, alongside vaccination, would result in an increase in malaria clinical cases and related deaths, surpassing baseline figures in the total population. Thus, even if the coverage (or distribution) of LLINs stays at the same average level as it was between 2020 and 2022, it is important to ensure that there is consistent net use. This can be promoted through SBCC.

To emphasize the importance of SBCC and to demonstrate its potential benefit for malaria prevention and control, Scenario 5 was included in our simulation. According to the previous three Benin DHS reports, there has been an increase in household population net accessibility (15% in 2006, 64% in 2011–2012, and 77% in 2017–2018) [32]. Similarly, the use of LLINs by household populations increased by 56% (from 14.7% in 2006 to 71.1% in 2018) [32]. However, the malaria indicators survey conducted in Benin in 2022 revealed that approximately 61% of the population use nets, lower than the 2018 reports [30]. Implementing high-quality strategic SBCC can enhance malaria prevention through campaigns aimed at raising awareness about the use of LLINs, the importance of seeking effective treatment, and other control measures [33].

In this regard, the primary behavioural objectives in Benin’s National Strategy for Malaria Social and Behavioural Change Communication 2021–2025 are to ensure that 90% of the population sleeps under LLINs every night, to promote early care-seeking and a 95% uptake of IPTp [3]. Given the critical role of SBCC in achieving these targets, we included Scenario 5 in our simulation to highlight its importance and potential impact on malaria prevention and control. Scenario 5, in which there is an increase in vaccine coverage compared to DTP3 levels and a 7.2% increase of the current effective LLIN coverage, resulted in a projected relative reduction of ∼62% in all outcomes compared to baseline. In addition to LLIN intervention, malaria vaccination combined with other control options, such as SMC, could potentially reduce the malaria burden beyond our current projections.

Several studies have evaluated the combination of malaria vaccination with chemoprevention and suggested this combination as a viable malaria control method. Chandramohan et al. [34], through an individually randomized controlled trial, revealed that the combination of vaccination and SMC resulted in a significant decrease in the incidence of malaria cases and deaths compared to implementing either intervention alone, highlighting the additional benefits of their combined use. Moreover, a mathematical modelling study demonstrated that adding seasonally targeted RTS,S to SMC would result in a greater reduction in clinical incidence compared to using SMC alone [35]. Therefore, future research should prioritize evaluating the impact and cost-effectiveness of combining the RTS,S malaria vaccine with SMC, considering variations in seasonality in the Benin context. Such studies are crucial for guiding policy decisions and optimizing resource allocation to maximize the reduction of malaria burden in children.

To the best of our knowledge this is the first national-level mathematical modelling study to estimate the impact of the RTS,S/AS01 malaria vaccine on malaria incidence in Benin. The findings of this study can provide valuable insights for policymakers and contribute to the growing body of evidence on the impact of RTS,S on malaria transmission in Benin.

Our study has several limitations. We use a country-level approach, which assumes the same incidence, vaccine coverage, and nets use at a national level. A detailed study at a sub-national level calibrated to local data is required to provide estimates of the vaccine impact at a sub-national level. We considered the primary series (3 doses) of the RTS,S/AS01 malaria vaccine. A fourth dose or any subsequent doses up to five doses delivered annually could potentially reduce the disease burden further than our projections. We considered the same coverage for LLIN use among both children and adults, but children are more likely to use nets than adults. Our approach was based on the malaria indicator survey, which indicated approximately 60% LLIN use (in both children and adults) in 2022, while the SBCC strategy aims to achieve 90% of the population sleeping under nets. Future research should consider this to provide a more accurate assessment of the impact of vaccination and LLIN use in children.

## Conclusion

This study employed a compartmental model to estimate the reduction in clinical malaria cases achieved by combining the RTS,S/AS01 vaccine with LLINs in Benin. Adopting childhood malaria vaccination at a coverage similar to the national DTP3 coverage levels, combined with the current use of insecticide-treated bed nets, is projected to result in a greater reduction in clinical malaria cases and deaths among children under five years old compared to using nets alone. However, the vaccine must be combined with a consistent distribution and use of nets to achieve substantial malaria burden alleviation, highlighting the need for continued relationships with donors.

## Data Availability

The data used in this study are open access and freely available from the respective sources.

## Author Contributions

Conceptualization, SA, SS, RH; Methodology, SA, SS, RH; Software, SA.; Formal analysis, SA.; Investigation, SA.; Resources, SS.; Writing—original draft, SA.; Writing— review & editing, S.S.,R.H.; Visualization, SA.; Supervision, S.S, RH.; Funding acquisition, S.S. All authors have read and agreed to the published version of the manuscript

