## Supplementary file for "Estimating the impact of the RTS,S/AS01 malaria vaccine in Benin: A mathematical modelling study"

### Mathematical model description

**Authors:** *Sena Alohoutade[1*], Rachel Hounsell[1,2], Codjo Dandonougbo[3], Rock Aikpon[3], Jules Degila[4], Sheetal Silal[1,2]*

[1] Modelling and Simulation Hub, Africa, Department of Statistical Sciences, University of Cape Town, Cape Town, South Africa

[2] Centre for Global Health, Nuffield Department of Medicine, Oxford University, Oxford, United Kingdom

[3] Programme National de Lutte contre le Paludisme, Ministere de la Sante, Benin

[4] Institut de Mathematiques et de Science Physiques, Dangbo, Benin

This document provides a description of the methodology, equations, parameters underlying the mathematical model and the supplementary material of the study.

The model is a deterministic age-structured dynamic compartmental model accounting for effective LLIN coverage and seasonality to estimate the impact of the RTS,S malaria vaccine on malaria cases and deaths in Benin. As the burden of malaria varies by age, the model considered the age structure of the population, determined by both the vaccination schedule and the age profile for the severity of the disease.

The model categorises the human population into three age groups: children from birth to six months, those aged six months to five years, and individuals aged five years and older. Susceptible individuals are exposed to infectious bites at a rate depending on mosquito density and infectivity. Following exposure ($E_i$), there is a latent period after which the infected individuals may experience an asymptomatic ($A_i$) episode of malaria according to their immunity level or develop clinical symptoms ($C_i$). The chance of symptomatic disease decreases with continued exposure due to the development of naturally acquired immunity. Asymptomatic individuals do not display symptoms but can transmit parasites to mosquitoes. However, asymptomatic individuals recover naturally after a period of time. A clinical episode, where individuals are symptomatic, can be uncomplicated ($C_i$) or progress to a severe malaria episode ($F_i$) which can result in death.

We assume that after recovery, individuals might be re-exposed and flow to the $L_i$ or $M_i$ compartment from which they might experience an asymptomatic or symptomatic episode of malaria at a different proportion from the first exposure.

We consider a natural mortality rate from each human compartments. However, malaria-attributable deaths are explicitly incorporated and considered in the severe stage of modelled infection. We assume that children age between groups at a rate $\zeta$.

We also assumed that susceptible, asymptomatic or recovered individuals at the age of six months may be vaccinated. Vaccinated individuals in the model may be infected and develop an asymptomatic or symptomatic episode of malaria. The vaccine effectiveness is defined as the relative reduction in clinical and severe malaria cases in the vaccinated compared to the unvaccinated population. Thus, after exposure the vaccinated children flow to their own compartments (Av, Cv) at a proportion that is different from the one of the unvaccinated children. Vaccinated individuals lose immunity due to waning after four years.

We included LLIN coverage and use in the scenarios to capture LLINs in the model.

Our model is based on the simple host-vector SEIR-SEI model with the extension to include several disease features.

The basic malaria model is defined as

$dS_m' = \mu_m M - ac\frac{I}{P}S_m - \mu_m S_m$ (1)

$dE_m' = ac\frac{I}{P}S_m - (\gamma_m + \mu_m)$ (2)

$dI_m' = \gamma_m E_m - \mu_m I_m$ (3)

$dS = \mu_h P - a\frac{M}{P}b\frac{I_m}{M}S + \rho R - \mu_h S$ (4)

$dE = a\frac{M}{P}b\frac{I_m}{M}S - (\gamma_h + \mu_h)E$ (5)

$dI = \gamma_h E - (r + \mu_h)I$ (6)

$dR = rI - (\rho + \mu_h)R$ (7)

Where M=$S_m$+ $E_m$+ $I_m$ and P=S+ E+ I+ R, the force of infection of the mosquito $\lambda_m$ is defined as $\lambda_m = ac\frac{I}{P}$ and the human force of infection is defined as $\lambda_h = ab\frac{I_m}{P}$

The model is then simplified by assuming that the mosquito population is at equilibrium. Assuming that $m = \frac{M}{P}$

The human force of infection is

$$\lambda_h = \frac{a^2bcm\frac{I}{P}}{(\mu_m+ac\frac{I}{P})}(\frac{\gamma_m}{\gamma_m+\mu_m})$$

The force of infection $\lambda_h$ has been modified in the model to account for LLINs.

We consider that a proportion of people that sleep under nets are protected from mosquito bites at a certain level depending on the effectiveness of the nets. We refer to this as effective LLIN coverage, which is comprised of the LLIN coverage, use and effectiveness.

In this regard (1-ITN) is the proportion of people that are not protected by bed nets, the force of infection is then

$$\lambda_h = \frac{(1-itn)*a^2bcm\frac{I}{P}}{(\mu_m+ac\frac{I}{P})}(\frac{\gamma_m}{\gamma_m+\mu_m})$$

The system is depicted in Figure 1 and described by the following set of ordinary differential equations with compartment descriptions in Table 1 and parameters description in Table 2.

**Key assumptions for each age group**

*The following are the assumptions that have been made for all the age group.*

- A clinical episode can be uncomplicated, leading to treatment, recovery, and a return to the susceptible compartment after immunity is lost.

- A clinical episode can progress to a severe episode, with some severe cases resulting in death.

- The potential to die from malaria infection occurs only when a patient experiences a severe episode of malaria.

*The following are the additional assumptions that applied for the second age group (6 to 60 months).*

- Only susceptible, asymptomatic, or recovered individuals at the age of 6 months can be vaccinated.

- Vaccinated individuals can still be exposed and develop either asymptomatic or symptomatic malaria, but they move to different compartments than unvaccinated individuals.

- The immunity from vaccination wanes after a certain period (assumed to be 4 years).

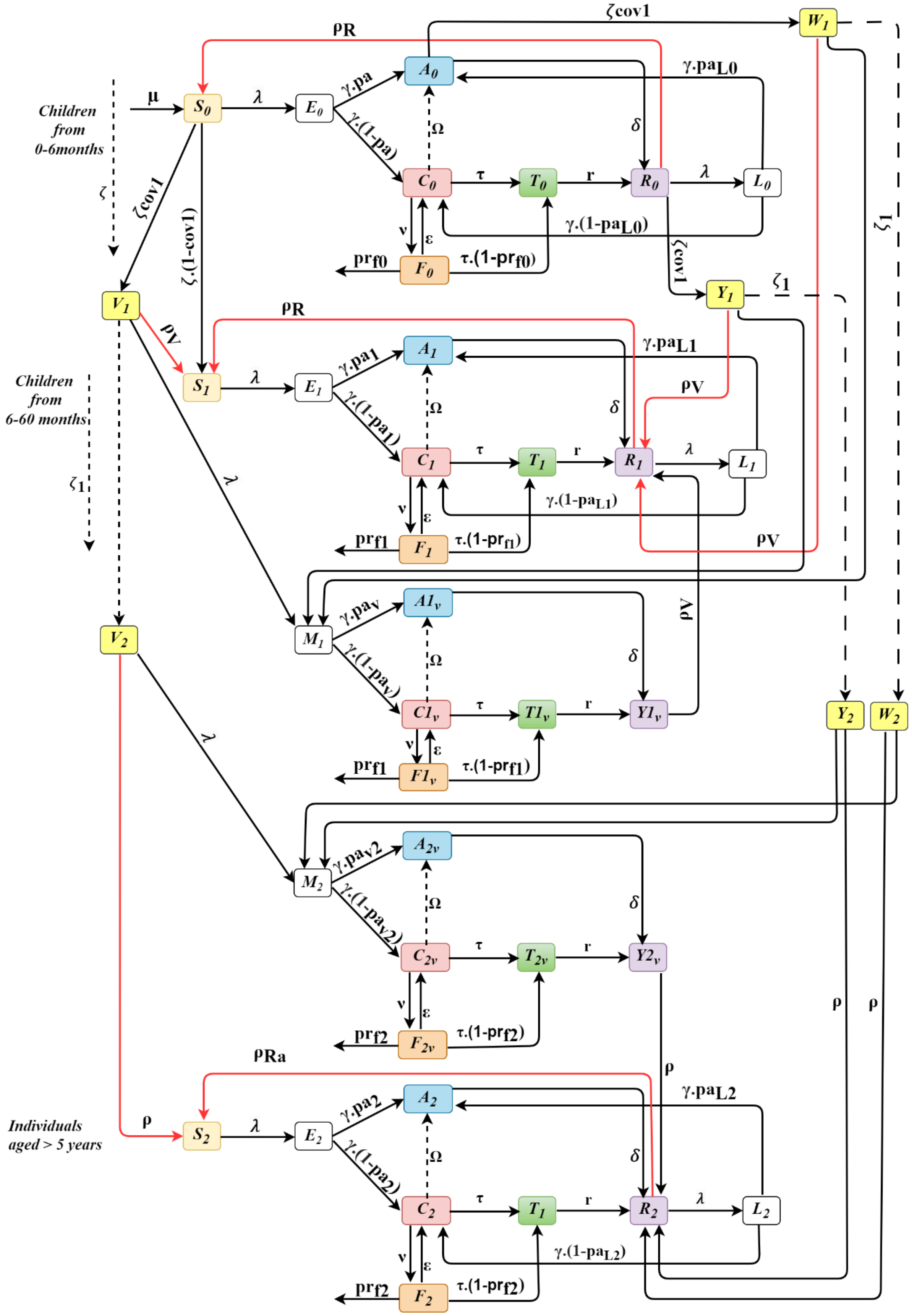


**Fig 1:** Compartmental model description with compartments $S_i$ (Susceptible) , $E_i$, $L_i$,$M_i$ (Exposed), $A_i$ (Asymptomatic), $C_i$ (Symptomatic), $T_i$ (Treated), $R_i$ (Recovered) and $V_i$, $W_i$,$Y_i$ (Vaccination compartments)

**Equations for children from birth to 6 months**

$$\frac{dS_0}{dt} = \mu_h P - \lambda S_o + \rho_R R_0 - \mu_h S_0 - \zeta.cov1.S_0 - \zeta.(1-cov1).S_0$$

$$\frac{dE_0}{dt} = \lambda S_0 - (\gamma_h + \mu_h)E_0 - \zeta E_0$$

$$\frac{dA_0}{dt} = \gamma_h.pa.E_0 - (\delta + \mu_h)A_0 + \omega.C_0 + \gamma_h.pa_{L0}.L_0 - \zeta.cov1.A_0 - \zeta.(1-cov1).A_0$$

$$\frac{dC_0}{dt} = \gamma_h.(1-pa)E_0 - (\tau + \mu_h)C_0 + \varepsilon.F_0 - (\nu + \omega)C_0 + \gamma_h.(1-pa_{L0})L_0 - \zeta C_0$$

$$\frac{dF_0}{dt} = \nu.C_0 - (\varepsilon + \mu_h).F_0 - \zeta.F_0 - \tau.\left(1 - pr_{f0}\right).F_0 - \tau.pr_{f_0}.F_0$$

$$\frac{dT_0}{dt} = \tau.C_0 - (r + \mu_h)T_0 + \tau.\left(1 - pr_{f0}\right).F_0 - \zeta T_0$$

$$\frac{dR_0}{dt} = rT_0 + \delta A_0 - (\rho_R + \mu_h + \lambda)R_0 - \zeta.cov1.R_0 - \zeta.(1-cov1).R_0$$

$$\frac{dL_0}{dt} = \lambda R_0 - (\gamma_h + \mu_h)L_0 - \zeta L_0$$

**Equations for children aged 6 months to 60 months (5 years)**

$$\frac{dS_1}{dt} = \zeta.(1-cov1).S_0 - \lambda S_1 + \rho_R R_1 + \rho_V V_1 - (\mu_h + \zeta_1)S_1$$

$$\frac{dE_1}{dt} = \zeta E_0 + \lambda S_1 - (\gamma_h + \mu_h)E_1 - \zeta_1 E_1$$

$$\frac{dA_1}{dt} = \zeta.(1-cov1).A_0 + \gamma_h.pa_1.E_1 - (\delta + \mu_h)A_1 + \omega.C_1 + \gamma_h.pa_{L1}.L_1 - \zeta_1 A_1$$

$$\frac{dC_1}{dt} = \zeta C_0 + \gamma_h.(1-pa_1)E_1 - (\tau + \mu_h).C_1 + \varepsilon.F_1 - (\omega + \nu)C_1 + \gamma_h.(1-pa_{L1})L_1 - \zeta_1 C_1$$

$$\frac{dF_1}{dt} = \zeta.F_0 + \nu.C_1 - (\varepsilon + \mu_h).F_1 - \zeta_1.F_1 - \tau.\left(1 - pr_{f_1}\right).F_1 - \tau.pr_{f_1}.F_1$$

$$\frac{dT_1}{dt} = \zeta T_0 + \tau.C_1 - (r + \mu_h)T_1 + \tau.\left(1 - pr_{f_1}\right).F_1 - \zeta_1.T_1$$

$$\frac{dR_1}{dt} = \zeta.cov1.R_0 + rT_1 + \delta A_1 - (\rho_R + \mu_h + \lambda).R_1 - \zeta_1.R_1 + \rho_Y.Y_1 + \rho_Y.Y_{1V} + \rho_W.W_1$$

$$\frac{dL_1}{dt} = \zeta L_0 + \lambda R_1 - (\gamma_h + \mu_h)L_1 - \zeta_1 L_1$$

$$\frac{dV_1}{dt} = \zeta.cov1.S_0 - \rho_V.V_1 - \lambda.V_1 - (\mu_h + \zeta_1).V_1$$

$$\frac{dW_1}{dt} = \zeta.cov1.A_0 - \rho_W.W_1 - \lambda.W_1 - (\mu_h + \zeta_1).W_1$$

$$\frac{dY_1}{dt} = \zeta.cov1.R_0 - \rho_Y.Y_1 - \lambda.Y_1 - (\mu_h + \zeta_1).Y_1$$

$$\frac{dM_1}{dt} = \lambda.V_1 + \lambda.W_1 + \lambda.Y_1 - (\gamma_h + \mu_h).M_1$$

$$\frac{dA_{1V}}{dt} = \gamma_h.paV.M_1 - (\delta + \mu_h).A_{1V} + \omega.C_{1V}$$

$$\frac{dC_{1V}}{dt} = \gamma_h.(1 - paV).M_1 - (\tau + \mu_h)C_{1V} + \varepsilon.F_{1V} - (\nu + \omega)C_{1V}$$

$$\frac{dF_{1V}}{dt} = \nu.C_{1V} - (\varepsilon + \mu_h).F_{1V} - \tau.\left(1 - pr_{f1}\right).F_{1V} - \tau.pr_{f1}.F_1$$

$$\frac{dT_{1V}}{dt} = \tau.C_{1V} - (r + \mu_h).T_{1V} + \tau.\left(1 - pr_{f1}\right).F_1$$

$$\frac{dY_{1V}}{dt} = r.T_{1V} + \delta.A_{1V} - \rho_Y.Y_{1V} - \mu_h.Y_{1V}$$

$$\frac{dV_2}{dt} = \zeta_1.V_1 - \rho.V_2 - \lambda.V_2 - \mu_h.V_2$$

$$\frac{dW_2}{dt} = \zeta_1.W_1 - \rho.W_2 - \lambda.W_2 - \mu_h.W_2$$

$$\frac{dY_2}{dt} = \zeta_1.Y_1 - \rho.Y_2 - \lambda.Y_2 - \mu_h.Y_2$$

$$\frac{dM_2}{dt} = \lambda.V_2 + \lambda.W_2 + \lambda.Y_2 - (\gamma_h + \mu_h).M_2$$

$$\frac{dA_{2V}}{dt} = \gamma_h.paV_2.M_2 - (\delta + \mu_h).A_{2V} + \omega.C_{2V}$$

$$\frac{dC_{2V}}{dt} = \gamma_h.(1 - paV_2).M_2 - (\tau + \mu_h).C_{2V} + \varepsilon.F_{2V} - (\nu + \omega)C_{2V}$$

$$\frac{dF_{2V}}{dt} = \nu.C_{2V} - (\varepsilon + \mu_h).F_{2V} - \tau.\left(1 - pr_{f_2}\right).F_{2V} - \tau.pr_{f2}.F_{2V}$$

$$\frac{dT_{2V}}{dt} = \tau.C_{2V} - (r + \mu_h).T_{2V} + \tau.\left(1 - pr_{f2}\right).F_{2V}$$

$$\frac{dY_{2V}}{dt} = r.T_{2V} + \delta.A_{2V} - \rho.Y_{2V} - \mu_h.Y_{2V}$$

**Equations for Individuals aged five years and older**

$$\frac{dS_2}{dt} = \zeta_1 S_1 - \lambda S_2 + \rho_R R_2 - \mu_h S_2 + \rho.V_2 + \rho_a.R_2$$

$$\frac{dE_2}{dt} = \zeta_1 E_1 + \lambda S_2 - (\gamma_h + \mu_h)E_2$$

$$\frac{dA_2}{dt} = \zeta_1 A_1 + \gamma_h.pa.E_2 - (\delta + \mu_h)A_2 + \omega.C_2 + \gamma_h.pa_{L2}.L_2$$

$$\frac{dC_2}{dt} = \zeta_1 C_1 + \gamma_h.(1 - pa_2)E_2 - (\tau + \mu_h).C_2 + \varepsilon_2.F_2 - (\omega + \nu_2)C_2 + \gamma_h.(1 - paL_2)L_2$$

$$\frac{dF_2}{dt} = \zeta_1.F_1 + \nu_2.C_2 - (\varepsilon_2 + \mu_h).F_2 - \tau.\left(1 - pr_{f2}\right).F_2 - \tau.pr_{f_2}.F_2$$

$$\frac{dT_2}{dt} = \zeta_1 T_1 + \tau_2.C_2 - (r_2 + \mu_h)\,T_2 + \tau.\left(1 - pr_{f_2}\right).F_2$$

$$\frac{dR_2}{dt} = \zeta_1 R_1 + rT_2 + \delta A_2 - (\rho_R + \mu_h + \lambda)R_2 + \rho.W_2 + \rho.Y_2 + \rho.Y_{2V} - \rho_a.R_2$$

$$\frac{dL_2}{dt} = \zeta_1 L_1 + \lambda R_2 - (\gamma_h + \mu_h)\,L_2$$

***Incorporating seasonality***

Seasonality was modelled by multiplying the force of infection of the model by a seasonality factor. The average monthly rainfall at a country level was standardized and fitted to trigonometric function, which was then used as a forcing function to account for seasonality.

The average monthly rainfall at a country level was downloaded as Climate Hazards Group InfraRed Precipitation with Station data (CHIRPS) for the period 2012 to 2023 from climateserv.servirglobal.net. We then use the manipulate package in R to conduct a simple calibration to ascertain the best value for parameters of the trigonometric function.

Seasonality factor = 1 + amp * cos(2*pi*(t - phase) / 365), the seasonality factor has been termed as "seas" in $\lambda_h$.

Resulting in $\lambda_h = \frac{seas*(1-itn)*a^2bcm\frac{I}{P}}{(\mu_m+ac\frac{I}{P})}\left(\frac{\gamma_m}{\gamma_m+\mu_m}\right)$

***Model fitting to data***

The mathematical model was calibrated to the estimated malaria case data of Benin to ensure that its predictions align as closely as possible with real-world observations. This calibration involved adjusting the model parameters to fit the estimated annual malaria case data sourced from the World Malaria Report over the years. This enhances its ability to capture malaria trends while accounting for factors such as intervention coverage and population-specific characteristics.

Figure 2 presents the calibration of the model against actual malaria case data from Benin. The plot compares the observed malaria cases (blue line) with the model's predictions (red dashed line) from 2016 to 2022. The data show a gradual increase in reported cases from 2016 to 2019, followed by a decline in 2020, and a subsequent slight rise until 2022. The model's predictions exhibit some variability, aligning more closely with the actual data in certain years, particularly in 2016, 2019, and 2022, suggesting reasonable model performance during those periods. However, the model overestimates malaria cases around 2020, predicting a peak that does not correspond to the observed trend. The model's predicted increase in cases from 2019 to 2020 contrasts with the actual decline in reported cases, which could be partially explained by the effects of the COVID-19 pandemic.

Due to concerns about contracting COVID-19, many individuals may have avoided seeking medical care, resulting in lower reported malaria cases, despite a potentially higher true incidence. Furthermore, under-reporting during 2020 may have occurred as health systems were overwhelmed by the pandemic, diverting resources and attention from other diseases, including malaria. This pandemic-related disruption may likely contribute to the discrepancies between the model's predictions and the observed data.

To improve the model's accuracy, collecting malaria case data at a finer temporal resolution, such as monthly or weekly, could enhance calibration efforts. More granular data would allow the model to better capture seasonal and short-term fluctuations in malaria transmission. In conclusion, while the model offers a reasonable estimate of malaria trends, its accuracy could be further improved through the integration of more detailed data.

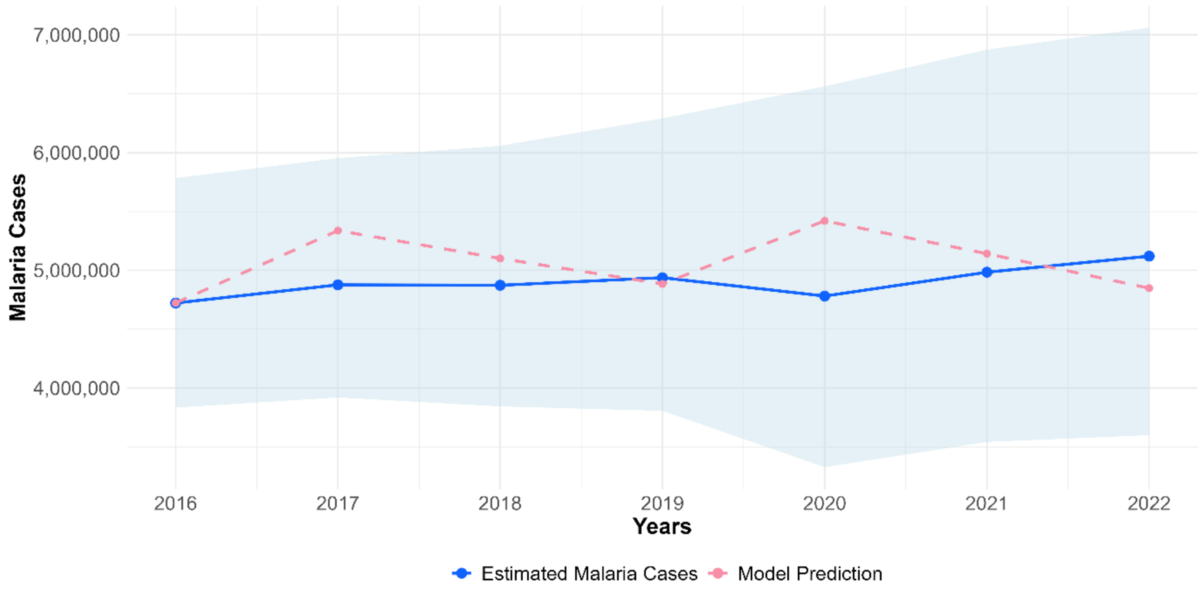


**Fig 2:** Calibration of the model to the estimated malaria cases in Benin (2016–2022)

*This plot shows the trends in estimated malaria cases (blue line) and model predictions (dashed pink line) from 2016 to 2022. The light blue shaded area represents the confidence intervals for the estimated malaria cases, indicating the range between the lower and upper values.*

Table 1 shows the various compartments, where i = 0, 1, 2, representing different age groups of the studied population

**Table 1. Description of compartments**

| Compartments | Description |
|---|---|
| $S_i$ | Susceptible population |
| $E_i$ | Exposed population |
| $A_i$ | Asymptomatic population |
| $C_i$ | Population with uncomplicated malaria |
| $F_i$ | Population with severe symptoms |
| $T_i$ | Treated population |
| $R_i$ | Recovered population |
| $L_i$ | Exposed population after recovery |
| $V_i$ | Vaccinated children from S compartment |
| $W_i$ | Vaccinated children from A compartment |

| | |
|---|---|
| $Y_i$ | Vaccinated children from R compartment |
| $M_i$ | Exposed children after being vaccinated |

**Table 2. Parameters table**

| **Symbol** | **Description** | **Value** | **Range** | **Source** |
|---|---|---|---|---|
| $1/\mu_h$ | Average life expectancy of the population | 62.22 | (62-63) | [1] |
| a | Human feeding rate per mosquito (per day) | 0.4 | (0.10-1) | [2,3] |
| b | Transmission efficiency from mosquito to human (per day) | 0.062 | (0.010-0.27) | [2,3] |
| c | Transmission efficiency human to mosquito (per day) | 0.4 | (0.072-0.64) | [2,3] |
| $\gamma_h$ | Rate of onset of infectiousness in humans (per day) | 0.071 | (0.066-0.08) | [3–6] |
| $\Omega$ | Duration of symptoms in untreated clinical infection (per day) | 5 | - | [4,7] |
| $\delta$ | Duration of symptoms in untreated asymptomatic infection (days) | 130 | (60,200) | [8] |
| $\tau$ | Treatment seeking rate (per day) | 0.33 | (0.175-0.38) | [9,10] |
| r | Recovery rate after antimalarial treatment (AL; per day) | 0.2 | (0.14-0.3) | [11,12] |
| $\rho_R$ | Duration of immunity in an adults (in year) | 4 | (3-5) | [4,13,14] |
| $\varepsilon$ | Duration of severe symptoms | 20 | - | assumption |
| $\nu$ | Duration of progress from clinical to severe infection | 7 | (3-10) | [15] |

| | | | | |
|---|---|---|---|---|
| pa | Proportion of asymptomatic children | 21 | na | [16,17] |
| VE | Vaccine effectiveness | 75 | (45-80) | [9,18,19] |
| itn_eff | Effectiveness of bednets | 44 | (37-46) | [20] |
| $\rho_V$ | Duration of immunity in vaccinated children | 4 | (3-5) | [18,19,21] |
| $\mu_m$ | Mosquito mortality rate (per day) | 0.066 | (0.05, 0.1) | [3,22] |
| $\gamma_m$ | Rate of onset of infectiousness in mosquitoes (per day) | 0.1 | (0.07,0.2) | [3,23] |
| amp | Amplitude of seasonal variation | 0.86 | (0,1) | *estimated from country data* |
| phase | Month of peak transmission | (6-7) | - | *estimated from country data* |
| $1/\zeta$ | Aging rate from birth to 6 months old | 6 | - | - |
| $1/\zeta_1$ | Aging rate from 6 months to 5-year-old | 54 | - | - |

***Estimation of uncertainty intervals for model outputs***

Uncertainty intervals were generated for all outcomes by performing 200 simulations per model scenario to account for parameter uncertainty. In each simulation, parameters were randomly sampled from their respective probability distributions (e.g., gamma, beta, triangular) within predefined lower and upper bounds. The choice of distributions was guided by the nature of each parameter's uncertainty, ensuring that the distribution parameters appropriately captured the expected range.

Gamma distributions were used for parameters that take strictly positive values, such as rates (e.g., recovery rate, treatment-seeking rate). Beta distributions were applied to parameters constrained within the [0,1] interval, such as proportions (e.g., the proportion of asymptomatic infections). Triangular distributions were employed for parameters with a known minimum, maximum, and most likely value, particularly when these values were informed by model calibration (e.g., human feeding rate per mosquito).

To efficiently sample from the multidimensional parameter space, the Latin Hypercube Sampling (LHS) method was used. Table 3 summarizes the distributions assigned to each parameter to represent uncertainty.

**Table 3. Distribution representing uncertainty regarding model parameters**

| Parameter | Distribution | Mean |
|---|---|---|
| Treatment seeking rate (per day) | Gamma (0.175,0.38) | 0.33 |
| Recovery rate after antimalarial treatment (per day) | Gamma(0.14,0.3) | 0.2 |
| Duration of symptoms in untreated clinical infection (per day) | Gamma(0.16,0.25) | 0.2 |
| Proportion of asymptomatic children | Beta(0.15,0.4) | 0.21 |
| Human feeding rate per mosquito (per day) | Triangular(0.38,0.45) | 0.4 |
| Rate of onset of infectiousness in humans (per day) | Triangular(0.066-0.08) | 0.071 |
| Rate of onset of infectiousness in mosquitoes (per day) | Gamma (0.07,0.2) | 0.1 |
| Duration of immunity in an adults (in year) | Gamma (3,5) | 4 |

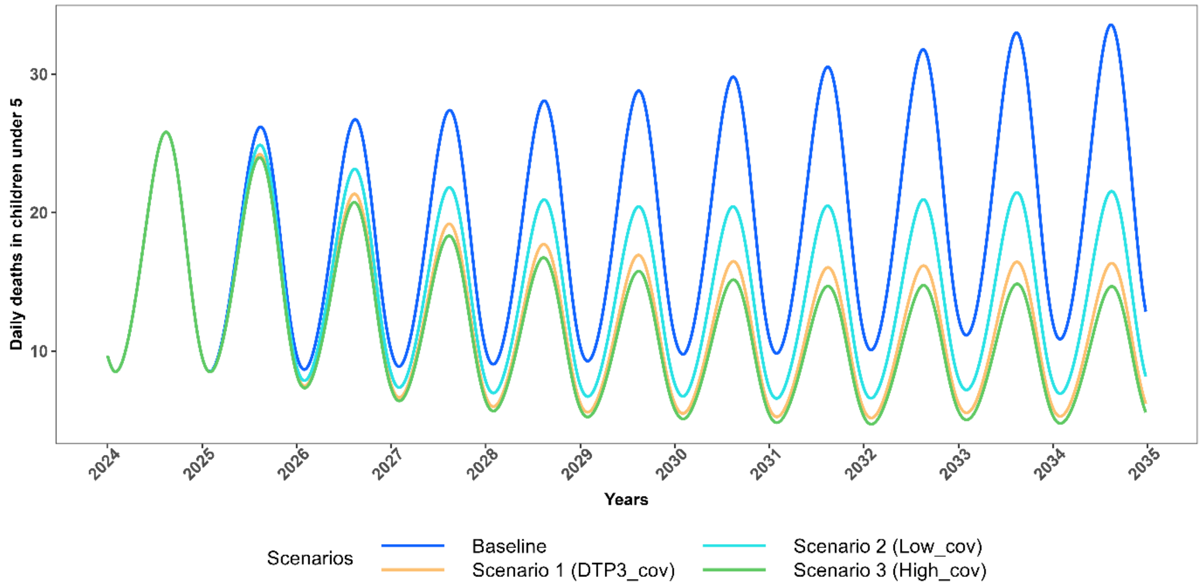


Fig 3: Projected trajectories of *P.falciparum* malaria daily deaths in CU5 for scenarios with no change in effective LLIN coverage (2025-2034)

*Trends in daily malaria related deaths in CU5 from 2024 to 2035, comparing intervention scenarios assuming no change in effective LLIN coverage (maintained at 43.2%). The Baseline scenario models 0% vaccination coverage (blue). Scenario 1 assumes 76% vaccination coverage with 43.2% effective LLIN coverage (orange); Scenario 2 assumes 50% vaccination coverage with 43.2% effective LLIN coverage (cyan); and Scenario 3 assumes 85% vaccination coverage with 43.2% effective LLIN coverage (green).*

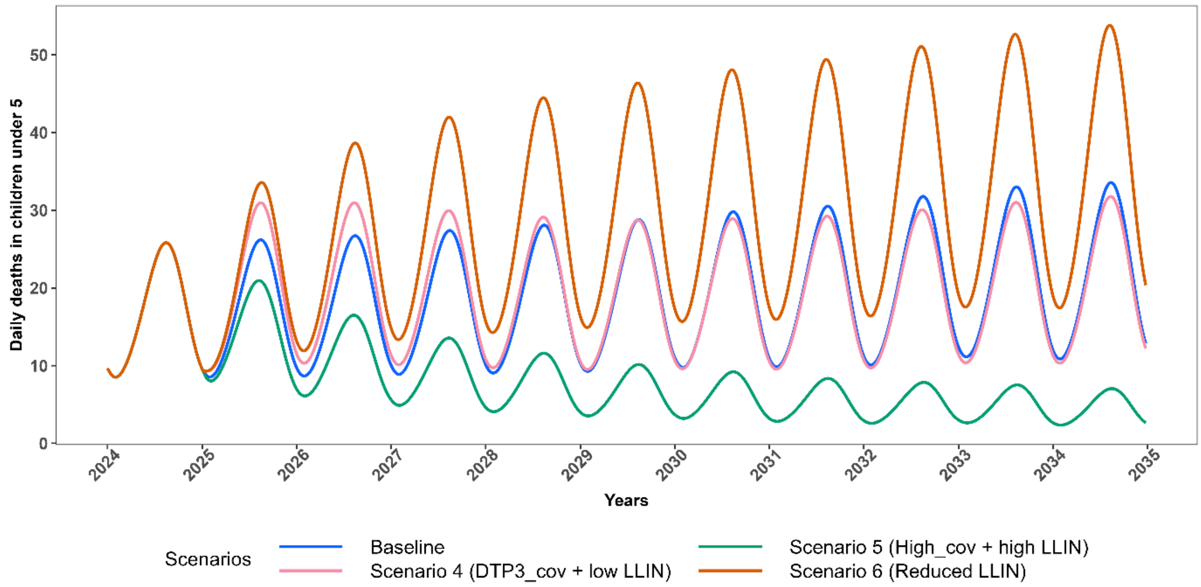


Fig 4: Projected trajectories of *P.falciparum* malaria daily deaths in CU5 for scenarios with change in effective LLIN coverage (2025-2034)

*Trends in daily malaria related deaths in CU5, comparing intervention scenarios involving changes in effective LLIN coverage to the baseline. The Baseline scenario assumes 0% vaccination coverage with 43.2% effective LLIN coverage (blue). Scenario 4 assumes 76% vaccination coverage with 28.8% effective LLIN coverage (pink); Scenario 5 assumes 85% vaccination coverage with 50.4% effective LLIN coverage (dark green); and Scenario 6 assumes 0% vaccination coverage with 28.8% effective LLIN coverage (dark orange).*

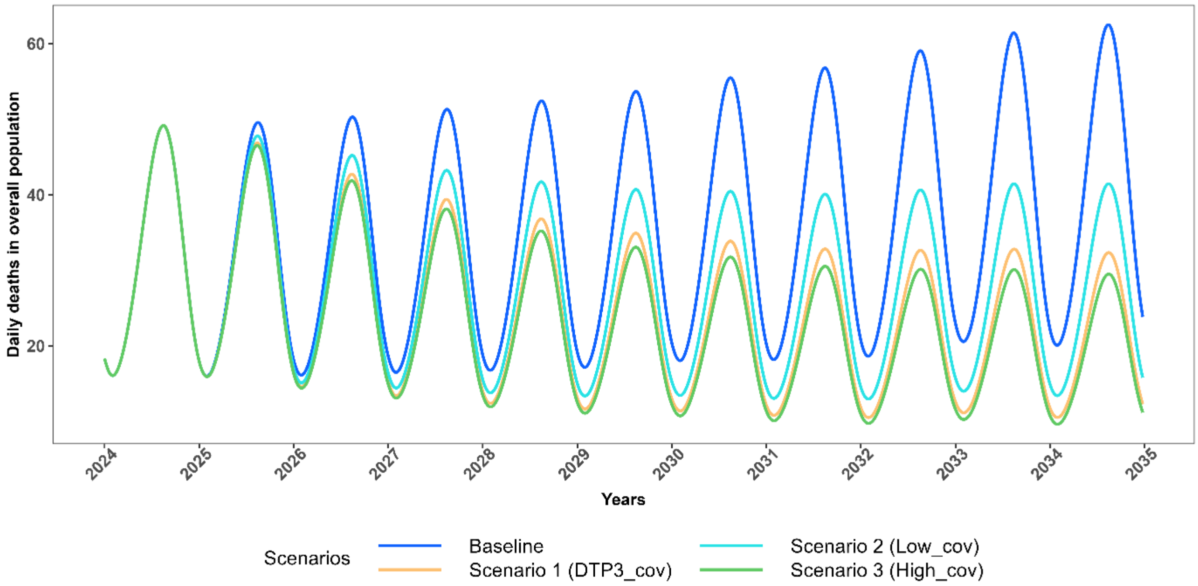


**Fig 5:** Projected trajectories of *P.falciparum* malaria daily deaths in the total population for scenarios with no change in effective LLIN coverage (2025-2034)

*Trends in daily malaria related deaths in the total population, comparing the projected impacts of intervention scenarios with no change in effective LLIN coverage to the baseline from 2024 to 2035. The Baseline scenario (blue), Scenario 1 (orange), Scenario 2 (cyan), and Scenario 3 (green) are depicted.*

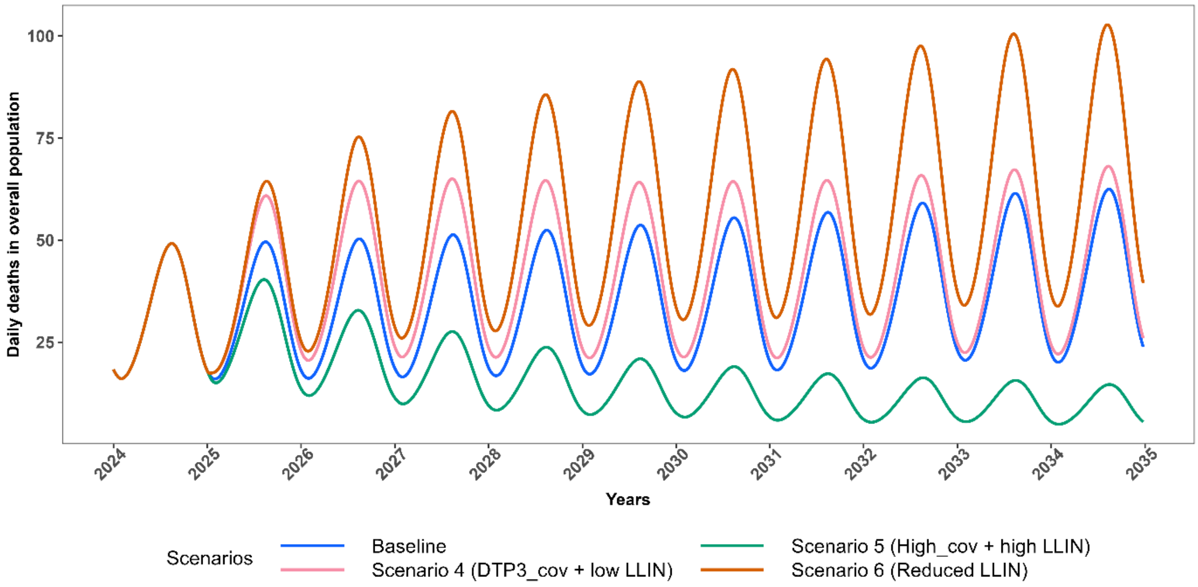


**Fig 6:** Projected trajectories of *P.falciparum* malaria daily deaths in the total population for scenarios with change in effective LLIN coverage (2025-2034)

*Trends in daily malaria related deaths in total population, comparing intervention scenarios involving changes in effective LLIN coverage to the baseline. The Baseline scenario assumes 0% vaccination coverage with 43.2% effective LLIN coverage (blue). Scenario 4 assumes 76% vaccination coverage with 28.8% effective LLIN coverage (pink); Scenario 5 assumes 85% vaccination coverage with 50.4% effective LLIN coverage (dark green); and Scenario 6 assumes 0% vaccination coverage with 28.8% effective LLIN coverage (dark orange).*

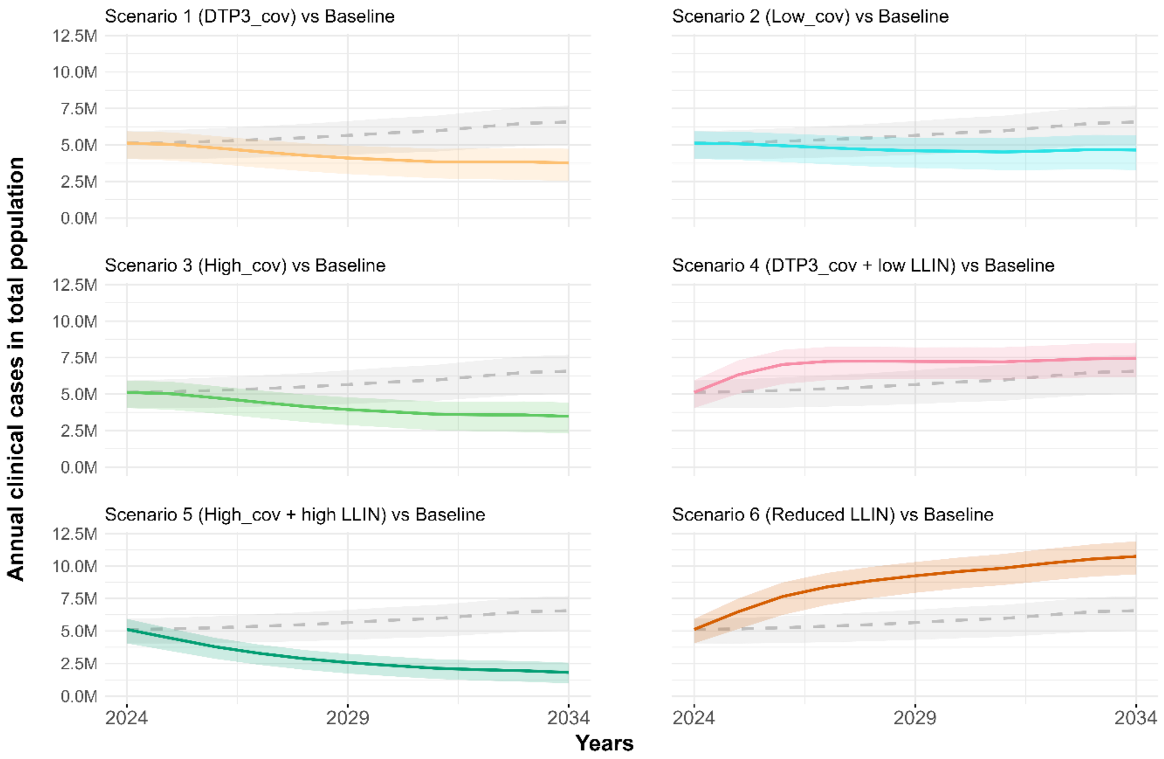


**Fig 7**: Projected trajectories of *P.falciparum* malaria annual cases in the overall population (2025-2034).

*The baseline is represented by a dashed grey line with a shaded grey confidence interval, indicating the range of uncertainty (90% uncertainty interval). Scenario 1 (orange), Scenario 2 (cyan), Scenario 3 (green), Scenario 4 (pink), Scenario 5 (dark green), and Scenario 6 (dark orange) show projections under different intervention assumptions. The Baseline scenario assumes 0% vaccination coverage with 43.2% effective LLIN coverage. Scenario 1 assumes 76% vaccination coverage with 43.2% effective LLIN coverage; Scenario 2 assumes 50% vaccination coverage with 43.2% LLIN coverage; and Scenario 3 assumes 85% vaccination coverage with 43.2% effective LLIN coverage.*

*Scenario 4 assumes 76% vaccination coverage with 28.8% effective LLIN coverage; Scenario 5 assumes 85% vaccination coverage with 50.4% LLIN coverage; and Scenario 6 assumes 0% vaccination coverage with 28.8% effective LLIN coverage. The solid lines represent the median estimates, while the shaded areas indicate the 90% uncertainty intervals (5th and 95th percentiles).*

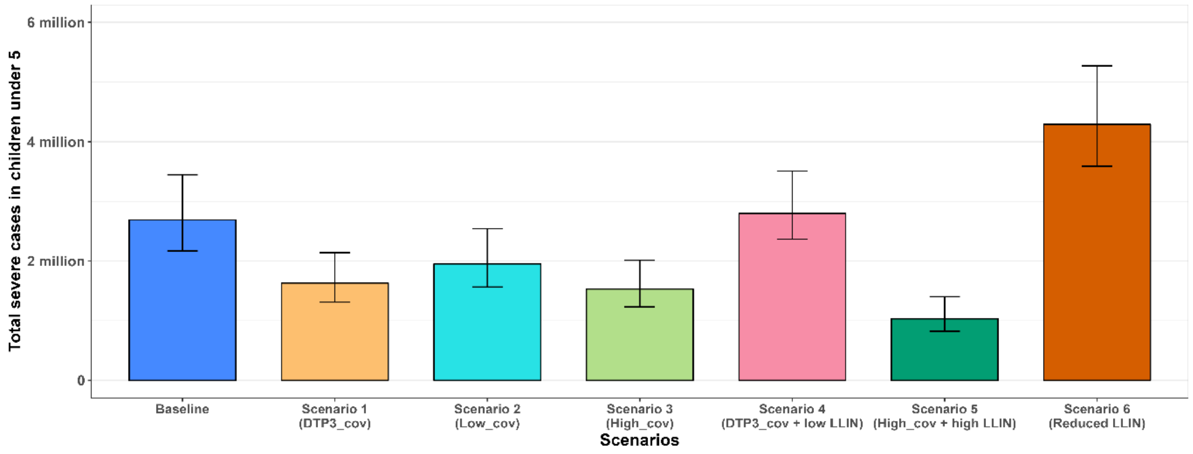


**Fig 8:** Total projected severe cases in CU5 (2025-2034)

*Bar graph showing the total clinical severe cases in CU5 for the baseline (blue) and six other scenarios: Scenario 1 (orange), Scenario 2 (cyan), Scenario 3 (green), and Scenario 4 (pink), Scenario 5 (dark green), and Scenario 6 (dark orange). The bars represent the 90% uncertainty intervals (5th and 95th percentiles) for each scenario.*

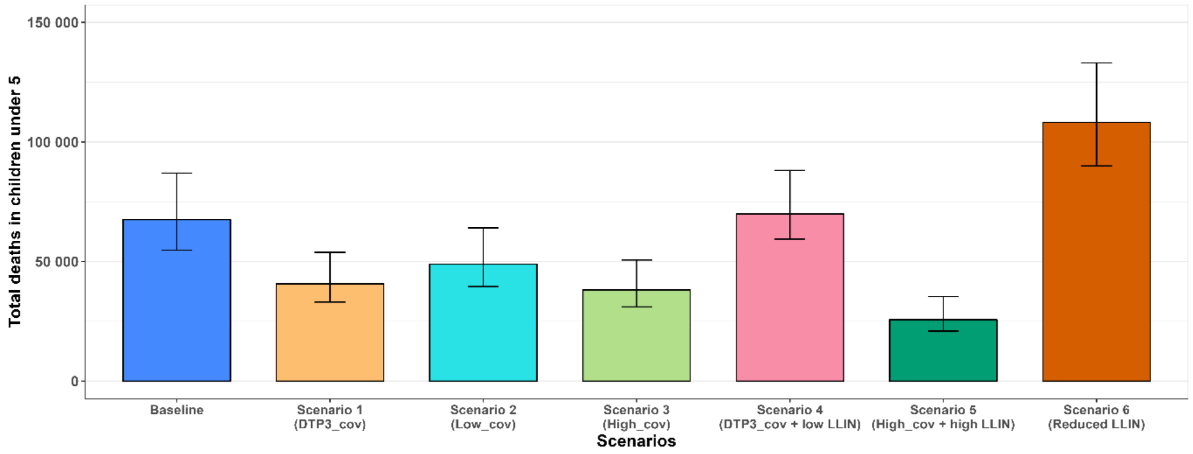


Fig 9: Total projected deaths in CU5 over 10 years (2025-2034)

*Bar graph showing the total deaths in CU5 for the baseline (blue) and six other scenarios: Scenario 1 (orange), Scenario 2 (cyan), Scenario 3 (green), and Scenario 4 (pink), Scenario 5 (dark green), and Scenario 6 (dark orange). The bars represent the 90% uncertainty intervals (5th and 95th percentiles) for each scenario.*

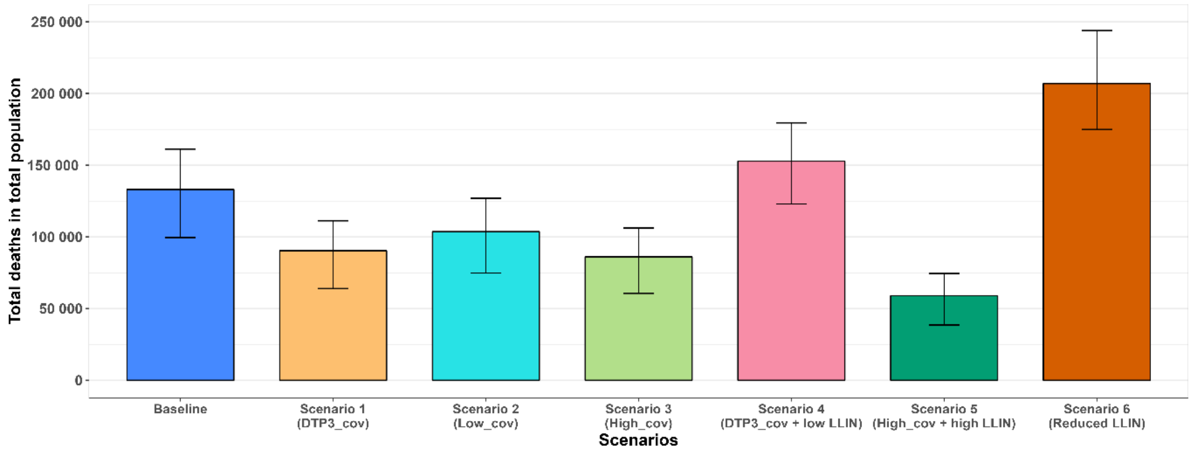


**Fig 10:** Total projected deaths in the total population over 10 years (2025-2034)

*Bar graph showing the total deaths in the total population for the baseline (blue) and six other scenarios: Scenario 1 (orange), Scenario 2 (cyan), Scenario 3 (green), and Scenario 4 (pink), Scenario 5 (dark green), and Scenario 6 (dark orange). The bars represent the 90% uncertainty intervals (5th and 95th percentiles) for each scenario.*

### Sensitivity analysis

A sensitivity analysis was conducted to evaluate the validity of our results and determine how changes in key parameters influence the projected impact on malaria cases over the 10-year period. This analysis considers two categories of parameters: the first set includes non-vaccine-related parameters, such as treatment-seeking period, net use, and net effectiveness, while the second set includes vaccine-related parameters that assess the potential influence of vaccination coverage, efficacy, and duration of protection on malaria outcomes. Figure 8 illustrates the outcome of the sensitivity analysis conducted on non-vaccine-related parameters.

On the first set of parameters, the sensitivity analysis revealed that net use and treatment-seeking period have the greatest impact on the number of malaria cases. A reduction in net use from 60% to 40% leads to an 83.72% increase in cases, while increasing net use to 70% reduces cases by 46.49%, highlighting the critical importance of net use. This strengthens the results of our simulation, confirming that increasing LLIN use can substantially reduce malaria cases.

Similarly, extending the treatment-seeking period from three to five days results in a 78.52% increase in cases, while reducing it to two days further decreases cases by 75.01%, emphasizing the need for prompt treatment to minimize cases. Net effectiveness also shows a moderate effect, with a 51.43% increase in cases when it is reduced, underscoring the importance of maintaining high-quality insecticide-treated nets. These results suggest that efforts to reduce malaria cases should focus on improving net use and encouraging early treatment-seeking behaviour.

Figure 9 presents a grid of heatmaps illustrating the percentage change in malaria cases based on varying vaccine coverage, vaccine effectiveness, and vaccine waning rates.

On the second set of parameters, the sensitivity analysis demonstrates that both vaccine coverage and vaccine waning are crucial in determining the overall impact of the malaria vaccine in CU5.

Lower coverage levels (e.g., 0.5) tend to be associated with less favourable outcomes, especially when combined with lower vaccine effectiveness and faster waning rates. Higher coverage (e.g., 0.85) leads to a more significant reduction in malaria cases, as seen by the green shades in the heatmaps. These findings reinforce the outcomes of our simulation, affirming that increasing vaccine coverage can significantly reduce the incidence of malaria cases.

In scenarios where the waning rate is slower (1/5 years), even with moderate coverage and effectiveness levels (e.g., 0.76), there is still a noticeable reduction in malaria cases. As the waning rate increases, the impact becomes less pronounced. For instance, in the 1/3 years column, lower effectiveness and coverage levels (bottom left of the grid) are associated with pink shades, indicating an increase in malaria cases due to the vaccine losing its effectiveness more quickly.

Vaccine effectiveness also shows a moderate effect, higher effectiveness (e.g., 0.85) leads to a reduction in malaria cases. However, lower effectiveness levels (e.g., 0.5) and in cases where vaccine coverage is also low, or the vaccine wanes quickly, the result is an increase in malaria cases, represented by the pink shades in the heatmaps.

Overall, high vaccine coverage (0.85) combined with high effectiveness (0.85) and a slower waning rate (1/5 years) produces the best outcomes, leading to a substantial reduction in malaria cases. Conversely, scenarios with low coverage, low effectiveness, or faster waning rates show limited impact or even an increase in malaria cases. Therefore, to maximize the impact of the vaccine, efforts should focus on achieving high coverage. This requires the allocation of adequate resources, the implementation of effective awareness campaigns, and the development of strategies to ensure broad distribution and acceptance of the vaccine.

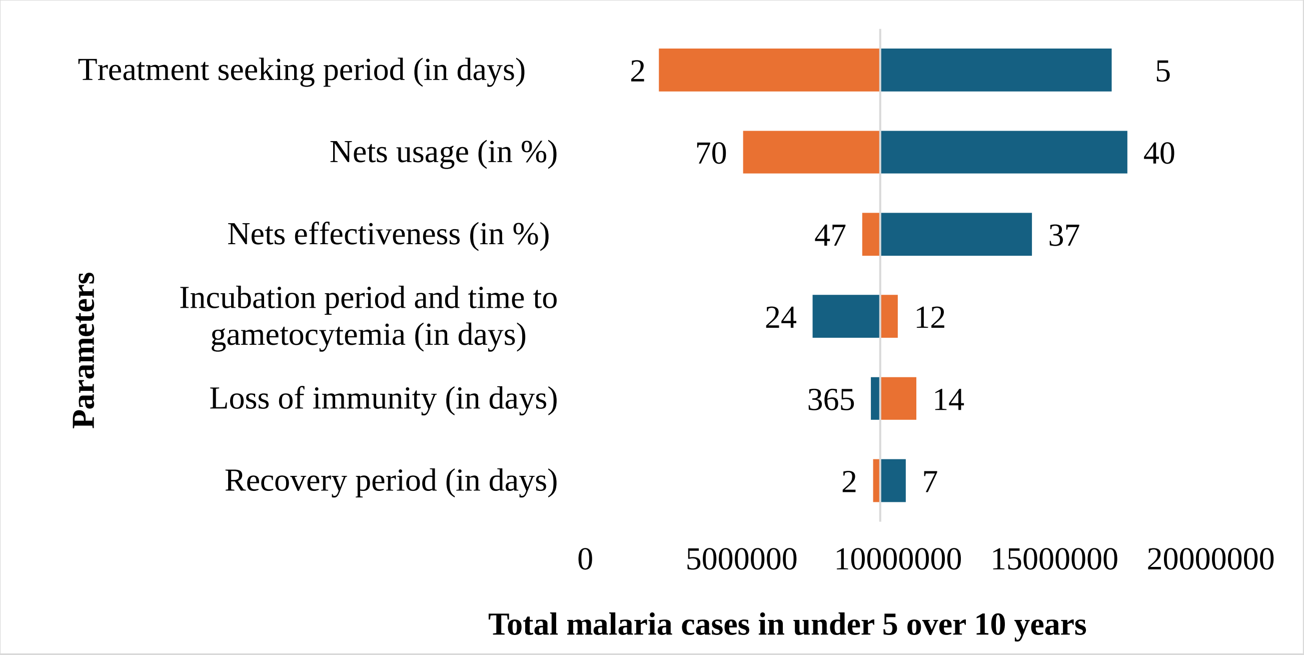


**Fig 11:** Sensitivity analysis conducted on non-vaccine-related parameters.

*The Tornado plot Shows sensitivity analysis of key parameters on malaria cases in children under 5: This plot highlights the impact of varying the recovery period, loss of immunity, incubation period, net effectiveness, net use, and treatment-seeking behaviour on malaria outcomes. A bar extending to the left indicates a reduction in malaria cases, while a bar extending to the right represents an increase*

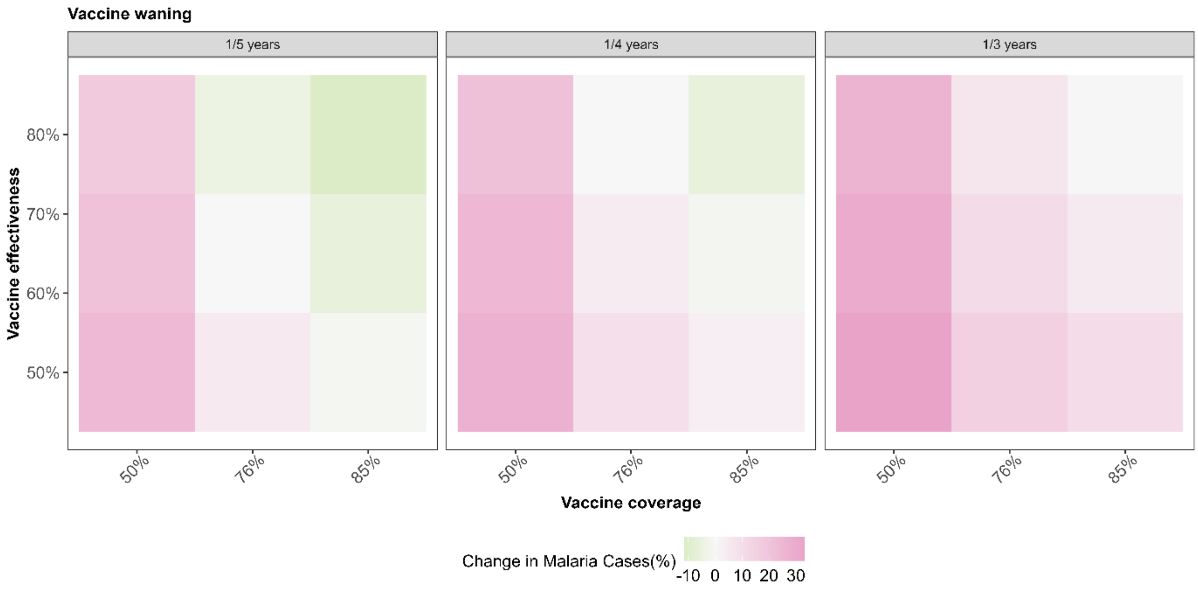


***Fig 12:*** Sensitivity analysis conducted on vaccine-related parameters

*This heatmap visualizes the percentage change in malaria cases among children under 5 years old, as a result of varying vaccine coverage (50%, 76%, 85%), vaccine effectiveness (50% to 80%), and vaccine waning rates (1/3, 1/4, 1/5 years). The color scale represents the magnitude of these changes, with green indicating a reduction in cases and pink indicating an increase.*

18. RTS,S Clinical Trials Partnership. Efficacy and safety of RTS,S/AS01 malaria vaccine with or without a booster dose in infants and children in Africa: final results of a phase

3, individually randomised, controlled trial. Lancet Lond Engl. 2015;386: 31–45. doi:10.1016/S0140-6736(15)60721-8
